# Midlife “Deaths of Despair” Trends in the US, Canada, and UK, 2001-2019: Is the US an Anomaly?

**DOI:** 10.1101/2022.10.10.22280916

**Authors:** Jennifer Beam Dowd, Colin Angus, Anna Zajacova, Andrea M. Tilstra

## Abstract

**Background:** Over the past decade, “deaths of despair” were strongly implicated in rising mid-life mortality in the US. Whether despair deaths and mid-life mortality trends are also changing in the peer countries such as the UK and Canada is not well known.

**Methods:** We compared all-cause and “despair” mortality trends at mid-life in the US, the UK (constituent nations England & Wales, Northern Ireland, and Scotland) and Canada from 2000-2019, using publicly available mortality data, stratified by three age groups (35-44, 45-54 and 55-64) and by sex. We examined trends in all-cause mortality and mortality by causes categorized as 1) suicides 2) alcohol-specific deaths 3) drug-related deaths. We employ several descriptive approaches to visually inspect age, period, and cohort trends in these causes of death.

**Results:** The US and Scotland both saw large increases and high absolute levels of drug-related deaths. The rest of the UK and Canada saw relative increases but much lower absolute levels in by comparison. Alcohol-specific deaths showed less consistent trends that did not track other “despair” causes, with older groups in Scotland seeing steep declines over time. Suicide deaths trended slowly upward in most countries.

**Conclusions:** In the UK, Scotland has suffered increases in drug-related mortality comparable to the US, while Canada and other UK constituent nations did not see dramatic increases. Alcohol-specific and suicide mortality generally follow different patterns to drug-related deaths across countries and over time, questioning the utility of a cohesive “deaths of despair” narrative.

## Introduction

The United States (US) has persistently lower life expectancy than its OECD peers, including 1.7-3.2 years below the United Kingdom (UK) and Canada in 2019 [1], two countries with a shared cultural and linguistic heritage. US life expectancy fell for three consecutive years starting in 2014, recovering slightly by 2019 prior to further declines in 2020 and 2021 during the COVID-19 pandemic [2, 3]. Explanations for stagnating life expectancy in the U.S and divergence from peer countries have highlighted increases in “deaths of despair” (i.e., deaths due to drug and alcohol poisoning, and suicide)[4, 5]. Yet, in the 2010’s there was evidence that life expectancy may also be stalling in England and Wales [6, 7] and Scotland [8] raising the question of whether these mortality dynamics reflect a broader phenomenon.

The US, the UK, and Canada have all documented rapid increases in drug-related mortality in recent years, with the US on the leading edge with an initial increase in deaths due to prescription opioids, followed by heroin and now fentanyl [9, 10]. Increases in opioid use in Canada may be related to exposure to US pharmaceutical promotional material through US media and direct marketing [11]. Within the UK, Scotland stands out with a dramatic rise in drug deaths (rising 450% since 1996) driven initially by prescription benzodiazepines and more recently by so-called ‘street’ benzodiazepines [12]. England and Wales have had a less extreme rise in drug-related deaths since 2012, driven by increasing numbers of opioid and cocaine-related deaths, and less noticeable increases due to benzodiazepines.^1^

Besides changes in the supply and marketing of drugs, increases in drug mortality may reflect demand side factors related to the loss of economic opportunity, particularly among socioeconomically disadvantaged groups [13, 14]. In Scotland, cohort trends in drug-related deaths and suicide were evident particularly in men from socioeconomically deprived areas coming of age in the 1980s [15, 16]. In the US and the UK, lack of economic opportunity in middle-aged cohorts may contribute to poorer mental health and subsequent coping behaviors [17]. In Canada, socioeconomic inequalities in health measured by educational attainment and household income have increased over time, but less is known about whether this has translated into despair related causes of death [18, 19].

This paper compares recent trends in mid-life “deaths of despair” and its component causes in the US to those in the UK and Canada from 2000-2019. We find that while increases in drug-related mortality were common across all nations, absolute levels are especially high in the US and Scotland. Overall, patterns in drug-related mortality were distinct from alcohol-specific and suicide deaths, questioning the utility of a cohesive “deaths of despair” framework.

## Data

Publicly available cause-specific mortality by age and sex were analysed for all countries. Because data are collected by separate statistical offices for the nations of the UK, and because of historically divergent mortality trends for Scotland, we estimate trends separately for UK nations, except for England and Wales whose data is released together. For England and Wales, we analyzed the 21^st^ Century Mortality dataset from 2001-2019,^2^ a publicly available resource that includes all registered deaths by cause as well as mid-year population counts by five-year age groups. Comparable data for Scotland is available from National Records of Scotland’s Vital Events Reference Tables^3^ and for Northern Ireland from The Registrar General’s Annual Reports.^4^ For Canada, death counts by cause of death were accessed from publicly available Statistics Canada Table 13-10-0392-01^5^. These records include all registered deaths in Canada each year from 2000-2019 by sex and 5-year age-group. US mortality data by year and single year of age come from CDC Wonder.^6^ Population counts by age and sex for all countries were obtained from the Human Mortality Database.

## Methods

We focus on mid-life mortality where increases in mortality have been seen in the U.S, specifically age groups 35-44, 45-54 and 55-64. We examine trends mortality by causes categorized as 1) suicides, 2) alcohol-specific deaths 3) drug-related deaths, and a combined 4) “deaths of despair” category as well as all-cause mortality. (ICD-10 codes available in supplemental Table S1). In order to mitigate the impact of random variation in the cause-and age-specific mortality rates we used a 1-dimensional P-spline approach for each country, cause, sex and calendar year to smooth the observed death rates over age [20]. The resulting single year of age-specific mortality rates were then age-standardised within each of the three broad age groups using the European Standard Population [21]. We visually inspected cohort vs. period trends using plots of the 3-year rolling averages within age-groups over time, supplemented with Lexis surfaces. Results are fully reproducible, and all data and code can be accessed here.

## Results

Full results for all age group, sex, country and cause groupings for all calendar years are shown in Appendix Tables S2a – S6b, with summary data for 2001, 2010 and 2019 presented in Tables 1a-b. Figure 1 shows all-cause mortality rates from 2001-2019 by age and sex group. Observed mortality patterns differ across nations and by sex. In Canada and England & Wales, mortality rates are low and decline consistently for most age-sex combinations, except for 35-44 year olds for which mortality trends are flat or increasing. Upticks in mortality were more noticeable in Scotland and the US in 35–44-year-olds, with steeper increases for males compared to females. In the US, all-cause mortality among males 35-44 reached a low point of 211.8/100,000 in 2012 before rising steadily to 257.3/100,000 in 2019 (Appendix Table S6a). Males aged 35-44 in Scotland followed a similar trend, with slightly higher absolute levels of mortality compared to US males. This youngest age group also showed signs of stalling improvements or slight increases in mortality in Canada, England & Wales, and Northern Ireland. Noticeable increases in overall mortality were also seen in Scotland for males and females ages 45-64, but not in the 55–64-year-old age group. The US was notable for its stagnation or increases in mortality across almost all age-sex groups.

**Table 1a.**
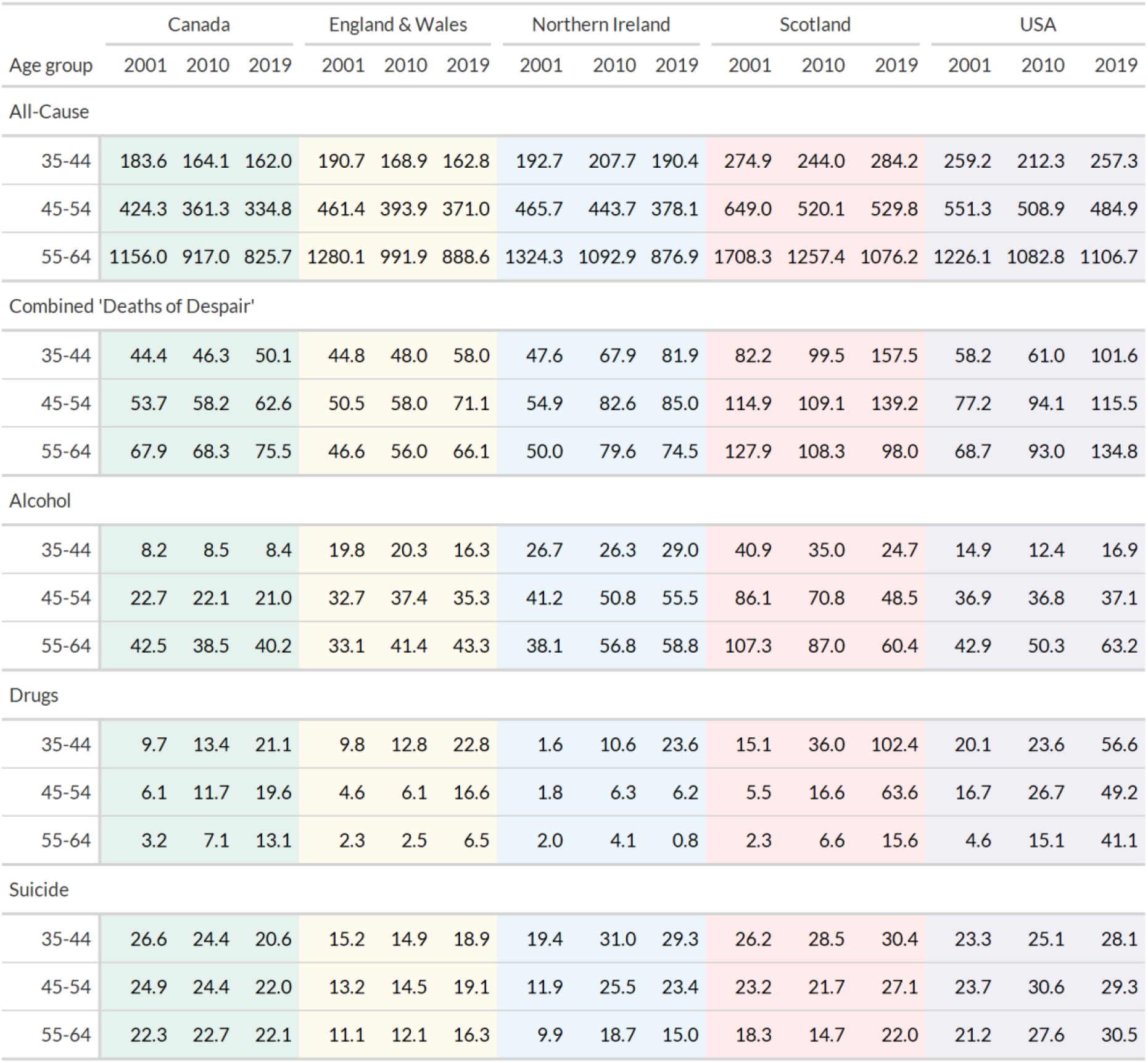
Male mortality rates (per 100,000 population) by cause and age group

**Table 1b.** Female mortality rates (per 100,000 population) by cause and age group

| Age group | Canada |  |  | England & Wales |  |  | Northern Ireland |  |  | Scotland |  |  | USA |  |  |
| --- | --- | --- | --- | --- | --- | --- | --- | --- | --- | --- | --- | --- | --- | --- | --- |
|  | 2001 | 2010 | 2019 | 2001 | 2010 | 2019 | 2001 | 2010 | 2019 | 2001 | 2010 | 2019 | 2001 | 2010 | 2019 |
| All-Cause |  |  |  |  |  |  |  |  |  |  |  |  |  |  |  |
| 35-44 | 112.1 | 100.2 | 94.6 | 117.5 | 104.8 | 98.7 | 119.4 | 122.3 | 115.0 | 149.2 | 133.4 | 153.4 | 147.6 | 128.1 | 142.8 |
| 45-54 | 268.6 | 235.9 | 212.6 | 302.5 | 260.4 | 243.0 | 310.2 | 292.7 | 274.0 | 387.9 | 328.6 | 341.1 | 319.8 | 311.7 | 295.1 |
| 55-64 | 685.8 | 586.7 | 530.0 | 802.5 | 641.2 | 587.8 | 809.9 | 704.9 | 639.7 | 1014.3 | 835.8 | 759.9 | 773.4 | 647.9 | 667.8 |
| Combined 'Deaths of Despair' |  |  |  |  |  |  |  |  |  |  |  |  |  |  |  |
| 35-44 | 15.9 | 19.1 | 19.0 | 16.8 | 18.3 | 23.2 | 20.1 | 26.3 | 32.3 | 35.6 | 37.4 | 61.7 | 22.0 | 27.7 | 40.5 |
| 45-54 | 20.0 | 24.3 | 24.1 | 23.0 | 24.6 | 30.5 | 29.4 | 39.6 | 43.3 | 48.4 | 46.4 | 64.7 | 26.3 | 42.8 | 51.2 |
| 55-64 | 24.1 | 26.7 | 28.9 | 23.7 | 25.4 | 29.4 | 24.6 | 36.3 | 42.5 | 49.9 | 44.9 | 53.3 | 24.1 | 35.1 | 53.8 |
| Alcohol |  |  |  |  |  |  |  |  |  |  |  |  |  |  |  |
| 35-44 | 4.0 | 4.2 | 4.3 | 9.6 | 10.2 | 9.7 | 15.0 | 13.9 | 17.1 | 19.9 | 17.9 | 12.8 | 6.4 | 5.5 | 8.6 |
| 45-54 | 8.6 | 8.5 | 9.5 | 16.4 | 17.3 | 18.2 | 22.9 | 25.9 | 29.9 | 36.7 | 32.2 | 28.4 | 12.1 | 14.6 | 17.7 |
| 55-64 | 14.7 | 13.2 | 16.4 | 17.7 | 19.6 | 20.5 | 20.8 | 26.1 | 32.7 | 41.3 | 35.6 | 37.5 | 15.8 | 17.3 | 26.7 |
| Drugs |  |  |  |  |  |  |  |  |  |  |  |  |  |  |  |
| 35-44 | 4.1 | 7.2 | 9.0 | 3.5 | 4.3 | 8.4 | 1.2 | 4.4 | 8.2 | 4.8 | 10.5 | 40.2 | 9.2 | 14.7 | 23.6 |
| 45-54 | 3.4 | 7.3 | 8.2 | 2.5 | 3.3 | 7.1 | 2.4 | 4.2 | 6.4 | 2.8 | 7.5 | 26.1 | 7.0 | 19.0 | 23.2 |
| 55-64 | 2.5 | 5.7 | 5.8 | 1.9 | 2.1 | 4.2 | 0.2 | 3.1 | 4.1 | 2.0 | 4.7 | 9.0 | 2.6 | 9.9 | 18.2 |
| Suicide |  |  |  |  |  |  |  |  |  |  |  |  |  |  |  |
| 35-44 | 7.8 | 7.6 | 5.7 | 3.8 | 3.8 | 5.2 | 3.9 | 7.9 | 7.0 | 10.9 | 8.9 | 8.7 | 6.5 | 7.4 | 8.2 |
| 45-54 | 8.0 | 8.4 | 6.5 | 4.2 | 4.0 | 5.3 | 4.1 | 9.5 | 7.0 | 9.0 | 6.7 | 10.2 | 7.2 | 9.2 | 10.3 |
| 55-64 | 6.9 | 7.8 | 6.7 | 4.1 | 3.7 | 4.7 | 3.6 | 7.1 | 5.8 | 6.6 | 4.6 | 6.8 | 5.7 | 7.9 | 8.9 |

**Figure 1.**
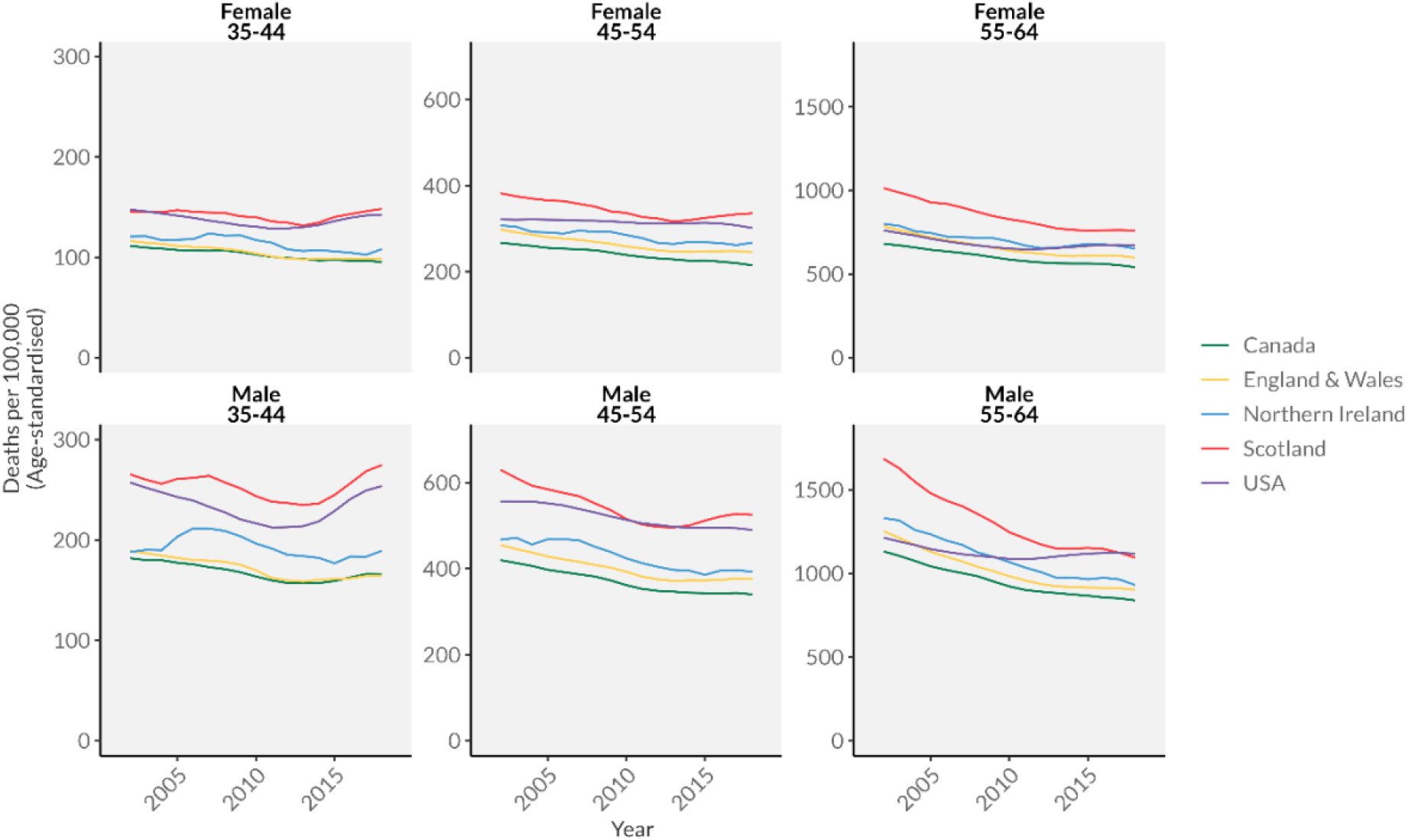
Midlife mortality from all causes by age and sex in Canada, the UK and the USA, 2001-2019

Figure 2. shows trends in “despair” deaths over the same period. The US and Scotland again stand out for substantially higher and largely increasing rates across the period. Males aged 35-44 show some of the worst trends among analyzed countries, with rates in the US and Scotland increasing almost 2-fold over the period. While the absolute increases for males in England and Wales and Northern Ireland are much smaller, they reflect large relative increases (for example a 41% increase in despair-related deaths for males ages 45-54 over this period in England and Wales). Canada is notable for its low levels and relatively small increases in deaths of despair, especially for women for whom rates hardly increase. While females in most nations saw only small increases in deaths of despair, the US stood out with a trend of high and rising for both males and females in all age groups.

**Figure 2.**
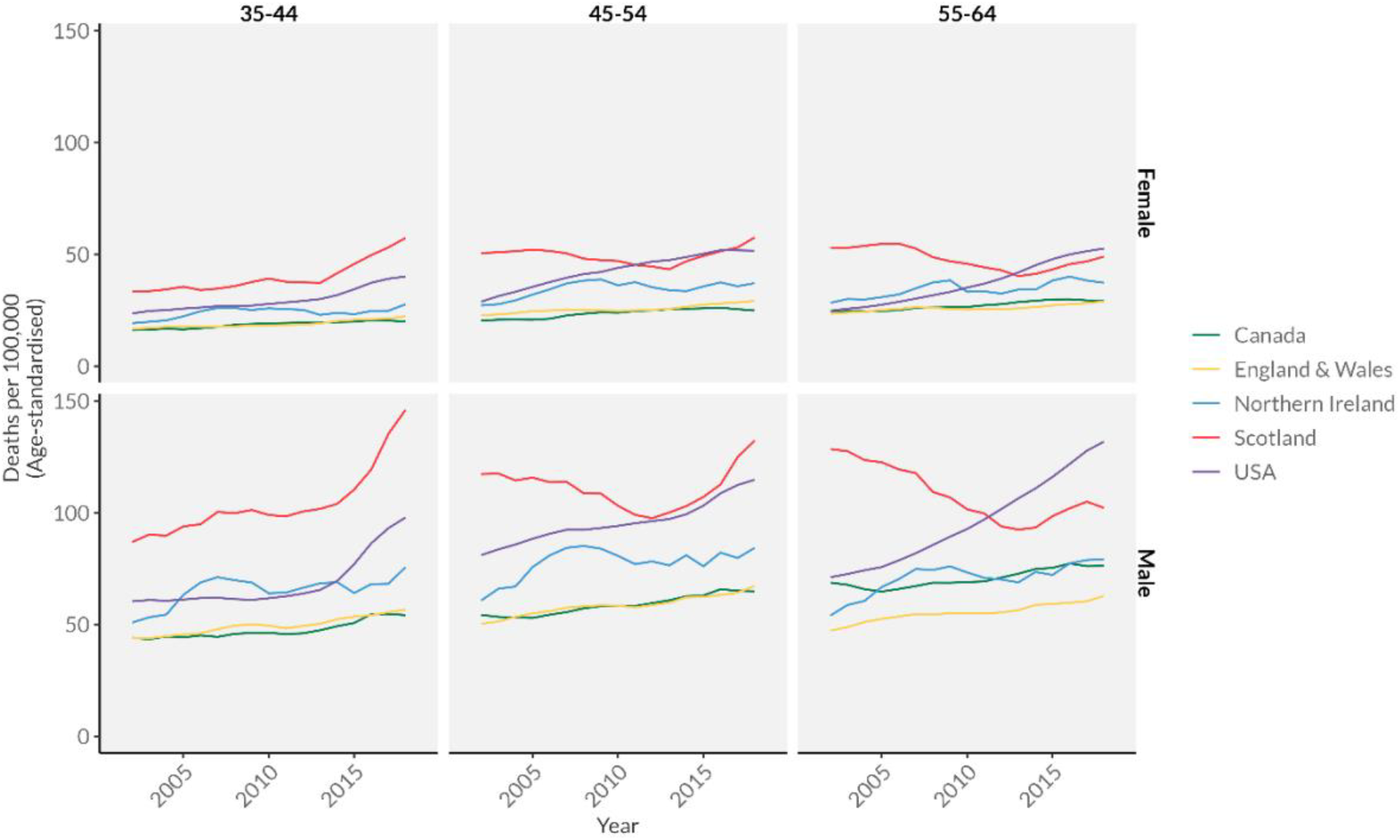
Combined mortality from ‘deaths of despair’ by age and sex in Canada, the UK and the USA, 2001-2019

Figures 2a-c break down these “despair” deaths into individual causes, where we see the salience of increasing drug-related mortality in the US and Scotland. The magnitude of the increases for males ages 35-44 in both countries was staggering (an almost 6-fold increase in Scotland from 15.1/100,000 to 102.3/100,000). In the US these increases are seen across all age groups, while in Scotland the increases are significantly less pronounced for 55–64-year-olds. While showing lower absolute levels, drug-related mortality also increased in Canada, England and Wales and Northern Ireland over the period, especially among younger males (for example an increase of 118% from 9.1/100,000 to 21.1/100,000 for Canadian males aged 35-44).

**Figure 2a.**
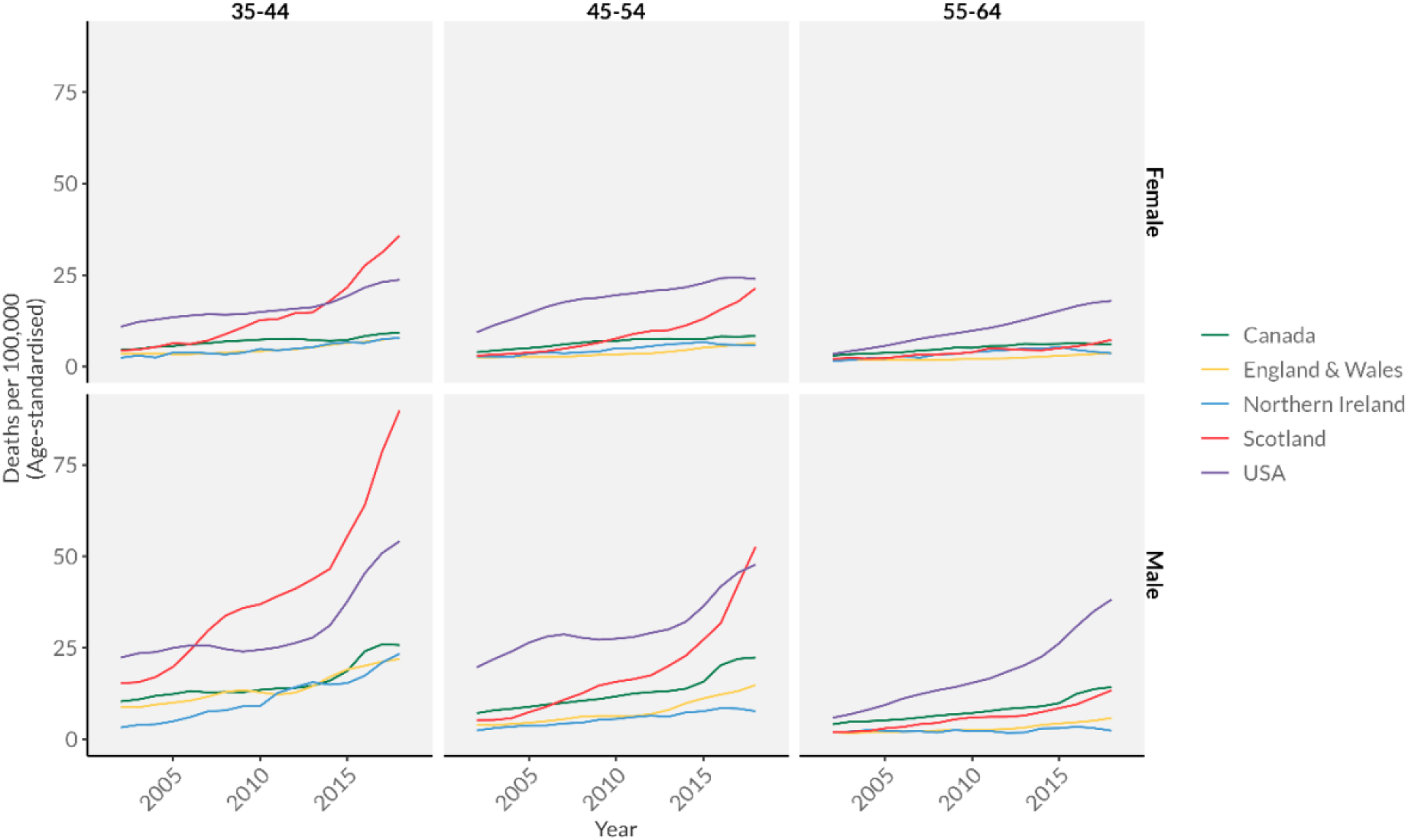
Drug-related mortality by age and sex in Canada, the UK and the USA, 2001-2019

**Figure 2b.**
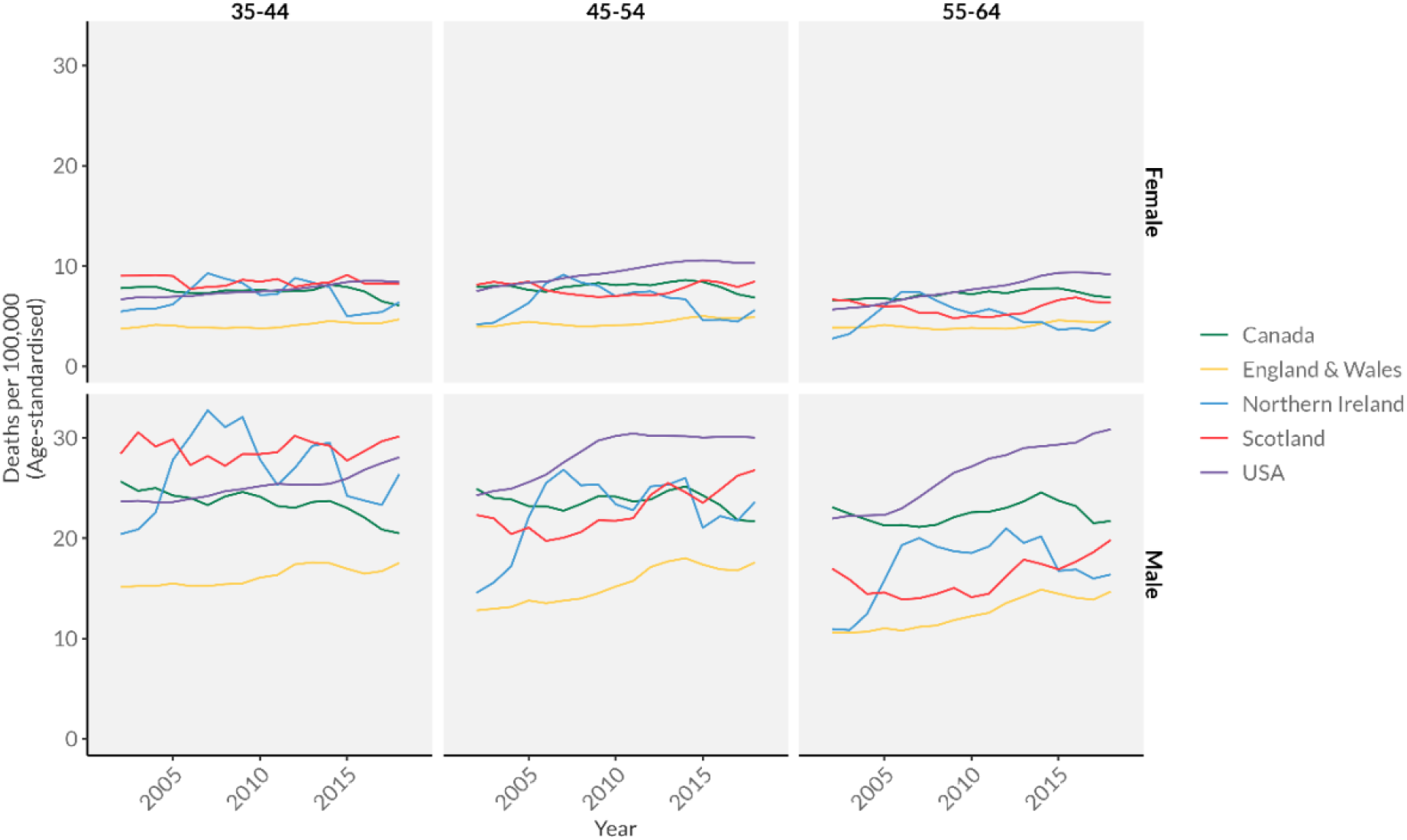
Suicide mortality by age and sex in Canada, the UK and the USA, 2001-2019

**Figure 2c.**
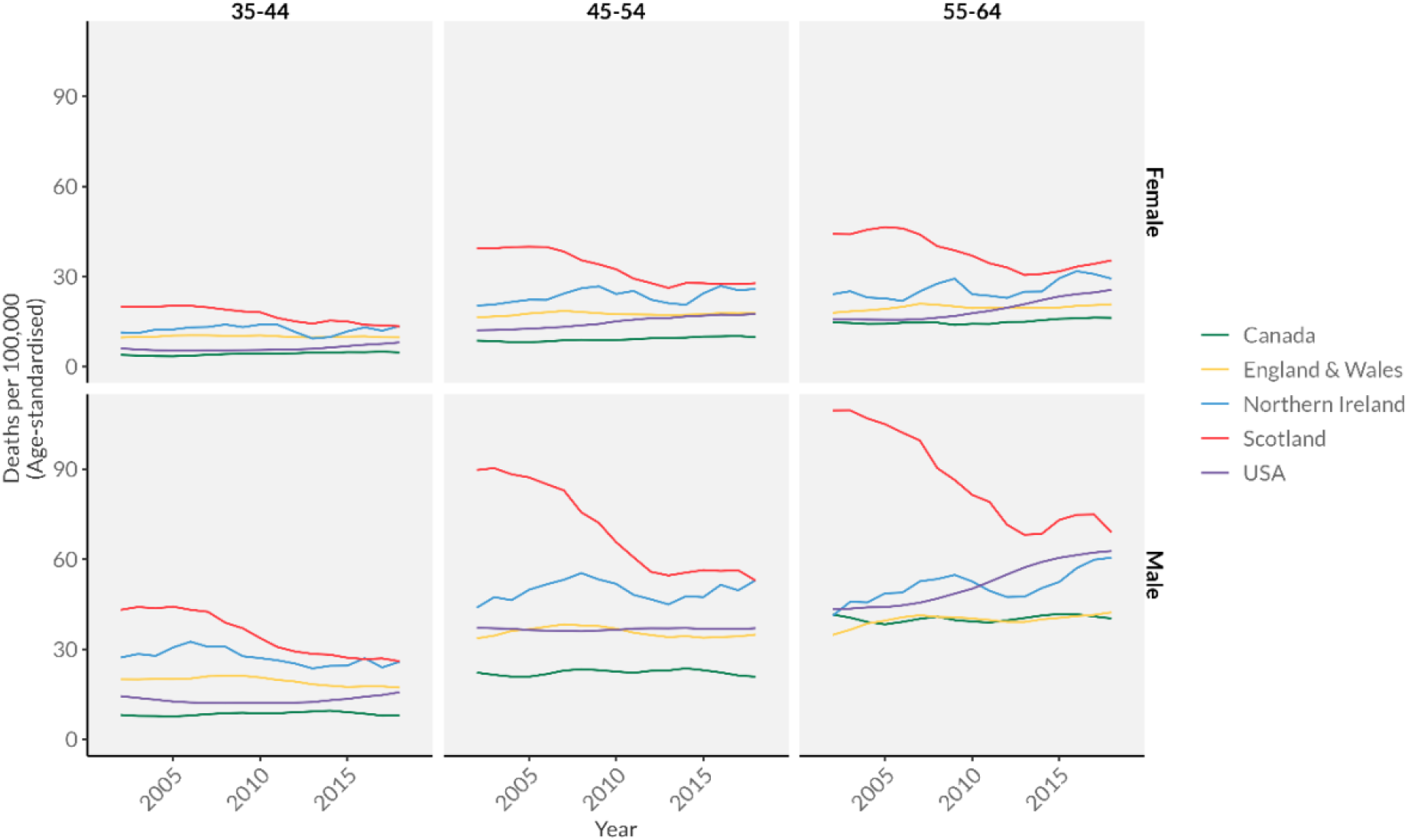
alcohol-specific mortality by age and sex

Suicide mortality saw more year-to-year variability, with evidence of small increases among males in England & Wales, Scotland, and the US. The magnitude of these changes is very small compared to changes in drug-related deaths. Suicide rates were higher for males in all nations. Canada consistently had the lowest levels of suicide mortality across all age-sex groups. The US generally had the highest levels of suicide deaths, except for males ages 35-44 where Scotland and Northern Ireland had higher levels. Alcohol-specific mortality across most nations was relatively stable, but with sharp declines over time in Scotland from previously very high levels, especially at the older ages. In contrast, increases in alcohol-specific mortality in the US were seen in ages 55-64, especially for men. Taken together, these results suggest that drug-related mortality is the primary cause of increases in overall “despair-related” mortality over this period, with small but perceptible increases in England & Wales, Canada, and Northern Ireland, and very pronounced increases in the US and Scotland.

**Figures S1a-c** show an alternative view of the same data to examine period vs. cohort trends. Here we look for “non-parallelism” in the age-specific mortality curves over time within each country [22]. If the age-specific curves are parallel to each other, this likely reflects the salience of period influences. Non-parallel age-specific trends suggest more importance for cohort effects (For a detailed discussion and simulations of these dynamics, see [23], pages 204-217). For overall deaths of despair, age-specific trends over time are mostly parallel for Canada, England & Wales, and Northern Ireland, suggesting period-based dynamics affecting all cohorts. The US and Scotland show some deviations from parallelism, with the youngest age group showing sharper increases than the older groups in Scotland, and the 55–64-year-olds in the US increasing more steeply (and earlier) than younger groups. For drug-related deaths, parallel trends are largely suggestive of period-based effects for Canada, England & Wales, and the US. Non-parallelism in Northern Ireland & Scotland suggests younger born cohorts have been hit harder by recent increases in drug deaths than the oldest cohort. For suicide, most trends are parallel suggesting period fluctuations, except for the US where the oldest age group has increased more steeply. Alcohol-specific mortality mostly follows period-based parallel trends, except for the US where again the oldest age group has increased over time. Cohort-driven declines in alcohol-related mortality were seen in Scotland, with the older groups declining steeply from high levels compared to lower and flatter trends in the youngest age group. Supplemental Figures S2a–b and show alternative visualizations for cohort patterns in mortality using Lexis surfaces. Overall, these visualizations highlight some limited cohort trends but also very strong period effects of increased drug deaths that affected most age groups and nations. In the US, increases in drug-related deaths affected all age groups over time, but sharper increases in alcohol-specific deaths and suicide were only seen in the oldest age group (earliest born cohort). This oldest group largely reflects the Baby Boom cohort (born 1946-1964) passing this 55-64 age group over the observed period.

## Discussion

Recent increases in mid-life mortality in the US set off a robust debate about the underlying causes of this concerning trend, with a strong focus on “deaths of despair.” One narrative emphasizes the role of cumulative disadvantage triggered by a worsening of labor market opportunities for white Americans with low levels of education, leading to psychological distress and unhealthy behavioral coping mechanisms [5, 24, 25]. A second hypothesis emphasizes the strong period trend in drug-related mortality due to changes in the availability of prescription opioids, heroin, and synthetic opioids (e.g., fentanyl)[26, 27]. While the first hypothesis implies that all “despair” deaths share a fundamental cause and move together, the latter argues that the trends in suicide and alcohol-specific mortality do not fit well within the overall deaths of despair narrative [28, 29].

Our study extends this work by comparing mid-life mortality trends in the US to Canada and the UK, two countries with shared sociocultural histories with the US but different economic, political and health care contexts. The UK has also experienced recent slowdowns in mortality improvements, raising the question of whether the US is the leading edge of more general mortality trends. Overall, we find evidence that the leveling off and increases in mid-life mortality seen in the US may be emerging more broadly, but the magnitude of these changes in other countries is muted. Scotland is the exception, with upward trends in drug-related deaths surpassing the high levels in the US and contributing to increases in overall mortality trends for males ages 35-54. While drug-related deaths have increased on a relative scale in the other UK nations and Canada, deaths from these causes are at much lower absolute levels than the US and Scotland. There is little evidence of significant trends in suicide or alcohol-specific deaths contributing to changes in all-cause mortality.

Our results suggest that increases in drug-related mortality are not necessarily unique to the US, with smaller increases also seen in Canada, England & Wales, and Northern Ireland, particularly among younger males. Scotland stands out as trending even higher than the US in drug-related mortality at younger ages, while the US saw parallel increases in drug-mortality across all age-ranges, suggesting a strong period effect. We found some evidence of cohort effects in alcohol-specific and suicide mortality in the US, with the oldest cohort trending up relative to younger cohorts. Suicide mortality rates were generally stable across countries for women, with slight increases for men and women in the US. Overall, with the exception of the oldest born (i.e. baby boom) male cohort in the US, we find that individual “deaths of despair” do not follow similar age and cohort patterns and thus may not reflect the same fundamental causes.

The fact that drug-related deaths have increased in other countries but not to the degree of the US is consistent with differences in prescribing practices and health care contexts. In the UK, there is no direct to consumer marketing by pharmaceutical companies, and the primary care system likely mitigates patients receiving prescriptions from multiple sources [30]. Nonetheless, prescribing for non-cancer pain did increase dramatically since the late 1990s when consensus statements supporting the use of opioids for the treatment of chronic pain [31, 32]. Despite this increase in overall opioid prescribing, a review of the UK Clinical Practice Research Datalink (CRPD) did not find an increase in the rate of diagnosed opioid use disorders in the UK between 2008 and 2012 [33]. While deaths from fentanyl in the UK have grown, they are still small relative to deaths from heroin or morphine, suggesting the context for drug prescribing and misuse may be different in the UK [34]. The dramatic rise in drug deaths in Scotland, with rates far outstripping those elsewhere else in Europe^7^ has arisen through a somewhat different mechanism to the US’ prescription opioid-driven crises. A combination of economic, social and political factors experienced by those living in deprived areas in the 1980s led to a rise in drug-related death rates in the 1990s [15, 16], driven by opioids such as heroin, often taken in combination with prescription benzodiazepines such as diazepam [35]. Although steps were taken to reduce the prescribing of benzodiazepines in the late 2000s, these were swiftly replaced by a range of ‘synethic benzodiazepines’ or ‘street benzos’ such as etizolam, which became widely and cheaply available through illicit markets [12]. The explosion in the use of street benzos has played a huge role in the rapid rise in Scottish drug deaths since 2012 when street benzos were implicated in 3% of all drug deaths compared to 66% in 2020, with concurrent use with opioids in particular the key factor – less than 1% of drug deaths in 2020 involved street benzos alone^8^. While concern over the opioid epidemic in Canada is rising, the absence of overall increases in mortality thus far has minimized public or scholarly discussion of a similar epidemic of “deaths of despair.”

## Conclusions

“Deaths of despair” due to alcohol, drug-overdoses, and suicide have been implicated in rising mid-life mortality in the US, but how these trends compare to peer countries is less well-known. We find that the US is unique in its large relative and absolute increases in drug-related mortality for all mid-life age-sex-groups, but that Scotland has seen a dramatic upward trend in drug deaths that now exceed the U.S. for younger age groups. The huge increase in drug mortality across all ages in the US from 2001-2019 is most consistent with period explanations, while alcohol and suicide upward trends are more focused in the oldest cohort. Overall, the trends in individual causes of despair deaths are not consistent across countries or age-groups, casting doubt on the underlying unity of a “deaths of despair” narrative.

## Data Availability

All data and analysis code are available on-line at: https://github.com/VictimOfMaths/DeathsOfDespair/tree/master/Paper

## Appendix Figures and Tables

**Figure S1.**
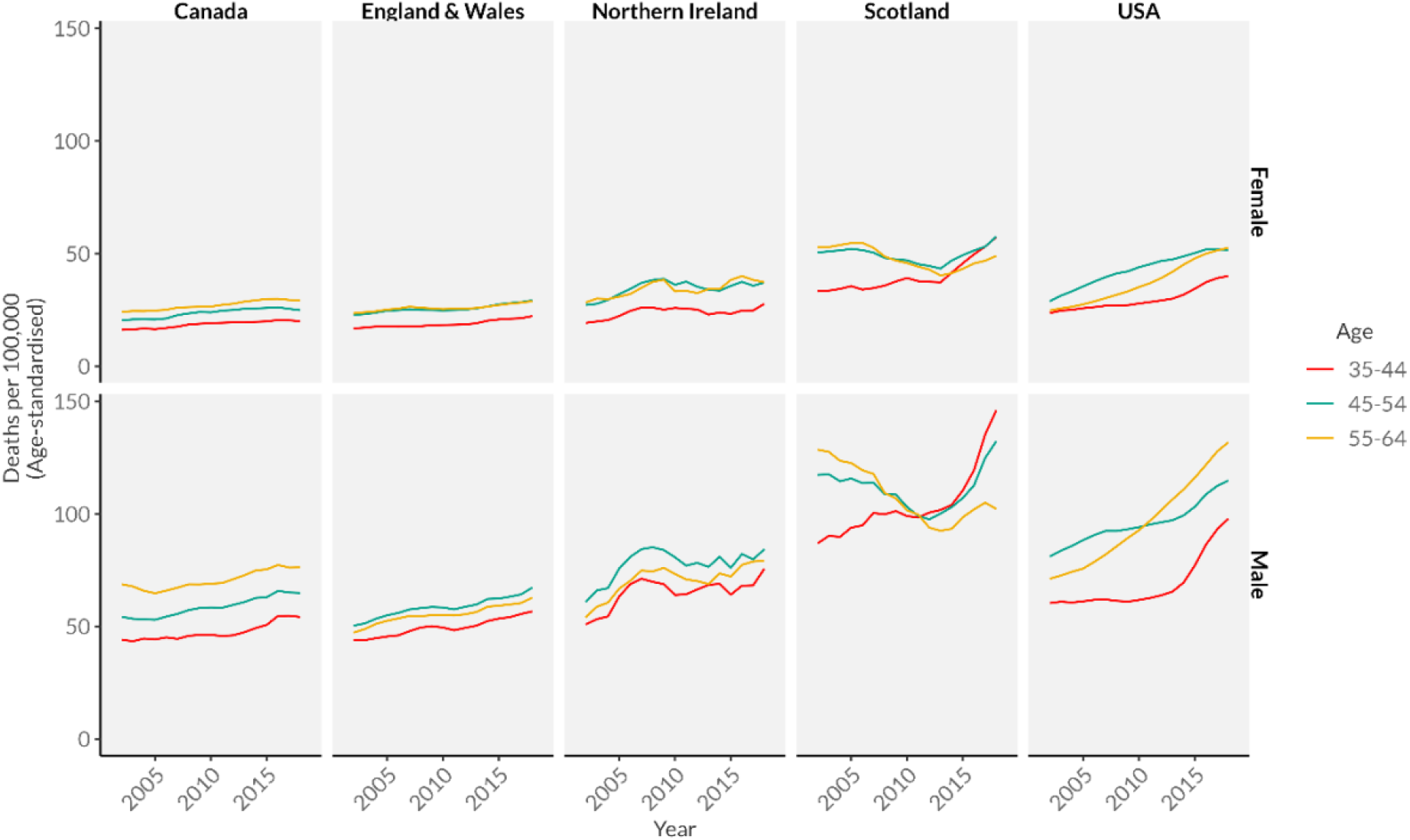
Mortality from combined ‘deaths of despair’ by age and sex in Canada, the UK and the US, 2001-2019

**Figure S1a.**
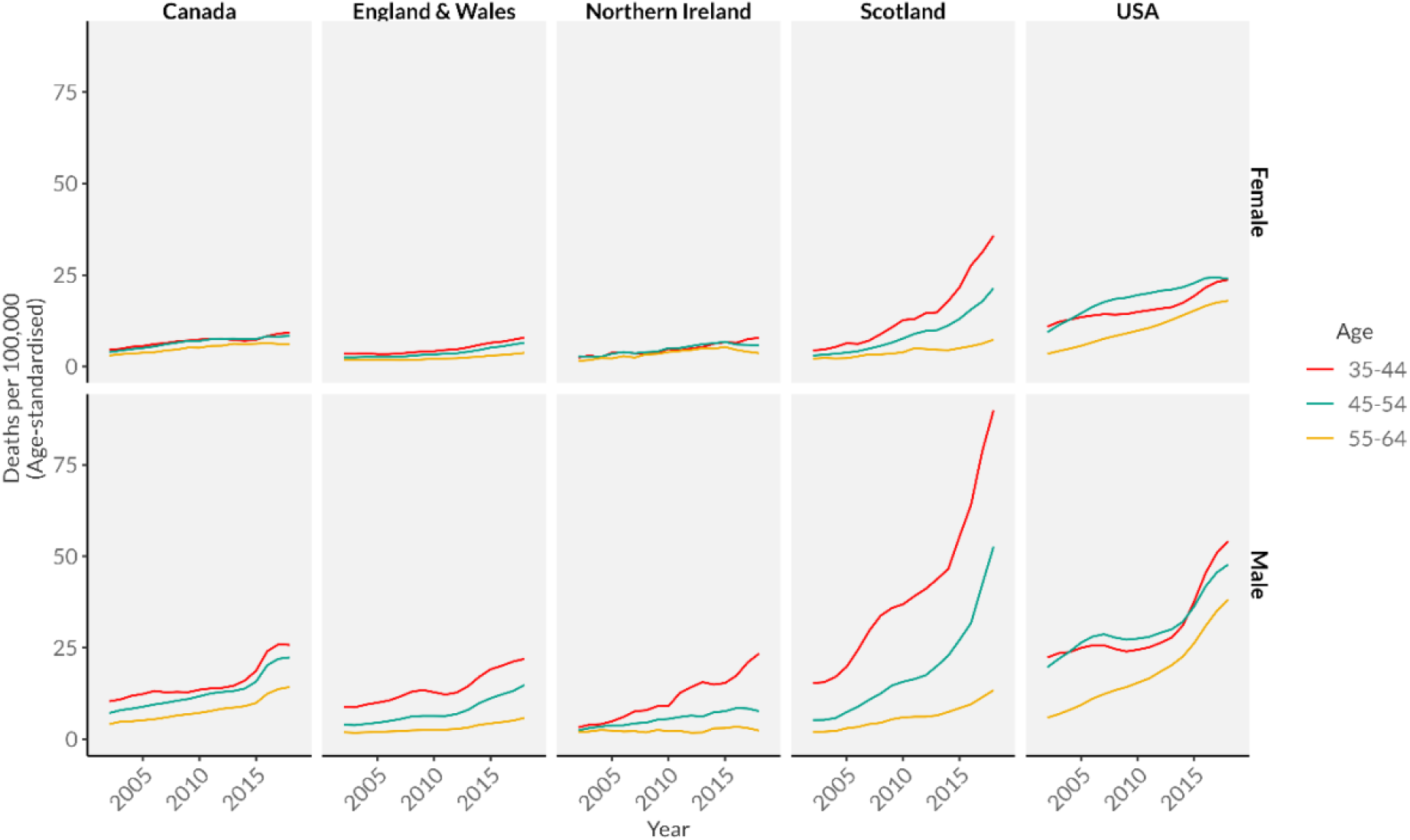
Drug-related mortality by age and sex in Canada, the UK and the US, 2001-2019

**Figure S1b.**
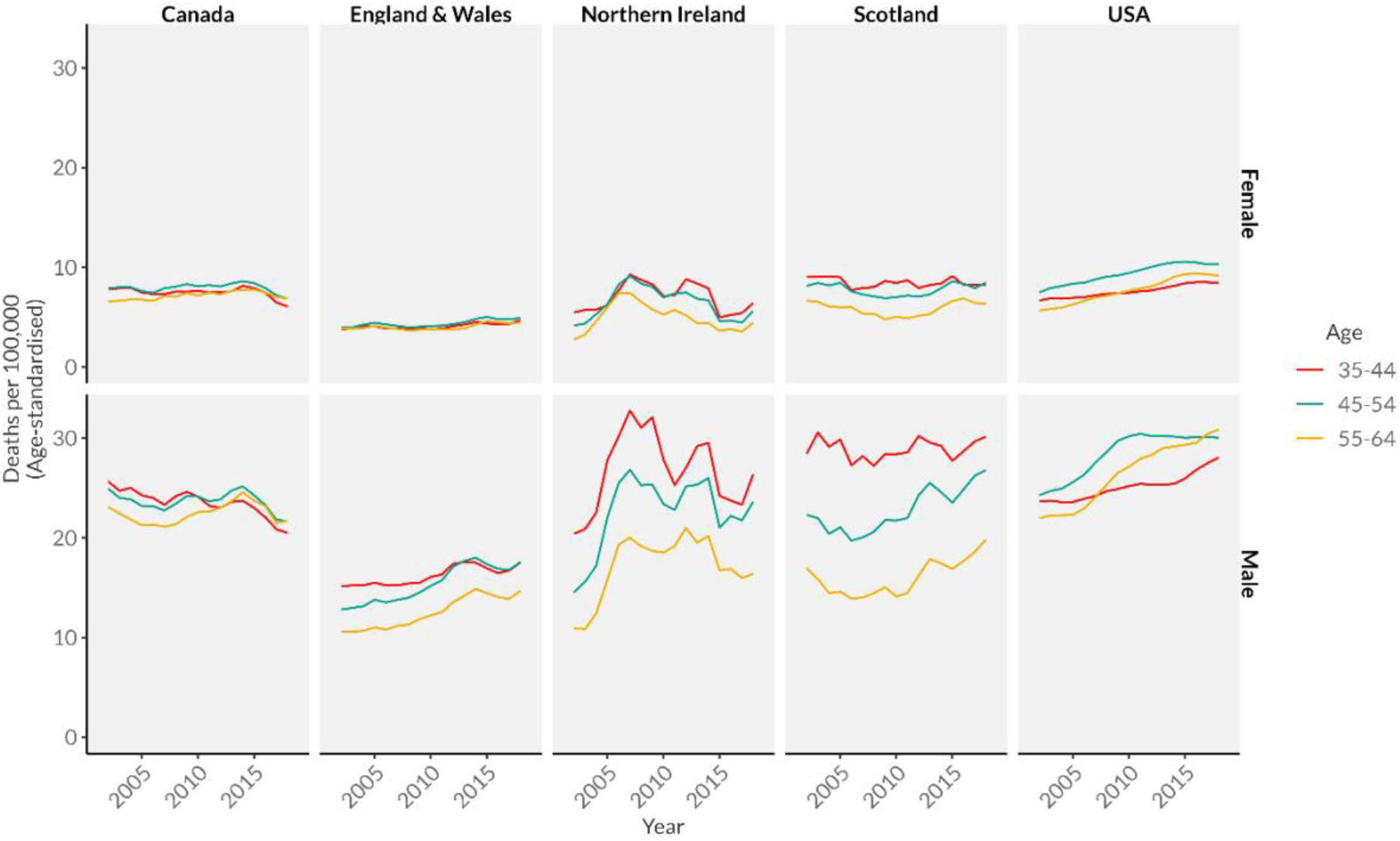
Suicide mortality by age and sex in Canada, the UK and the US, 2001-2019

**Figure S1c.**
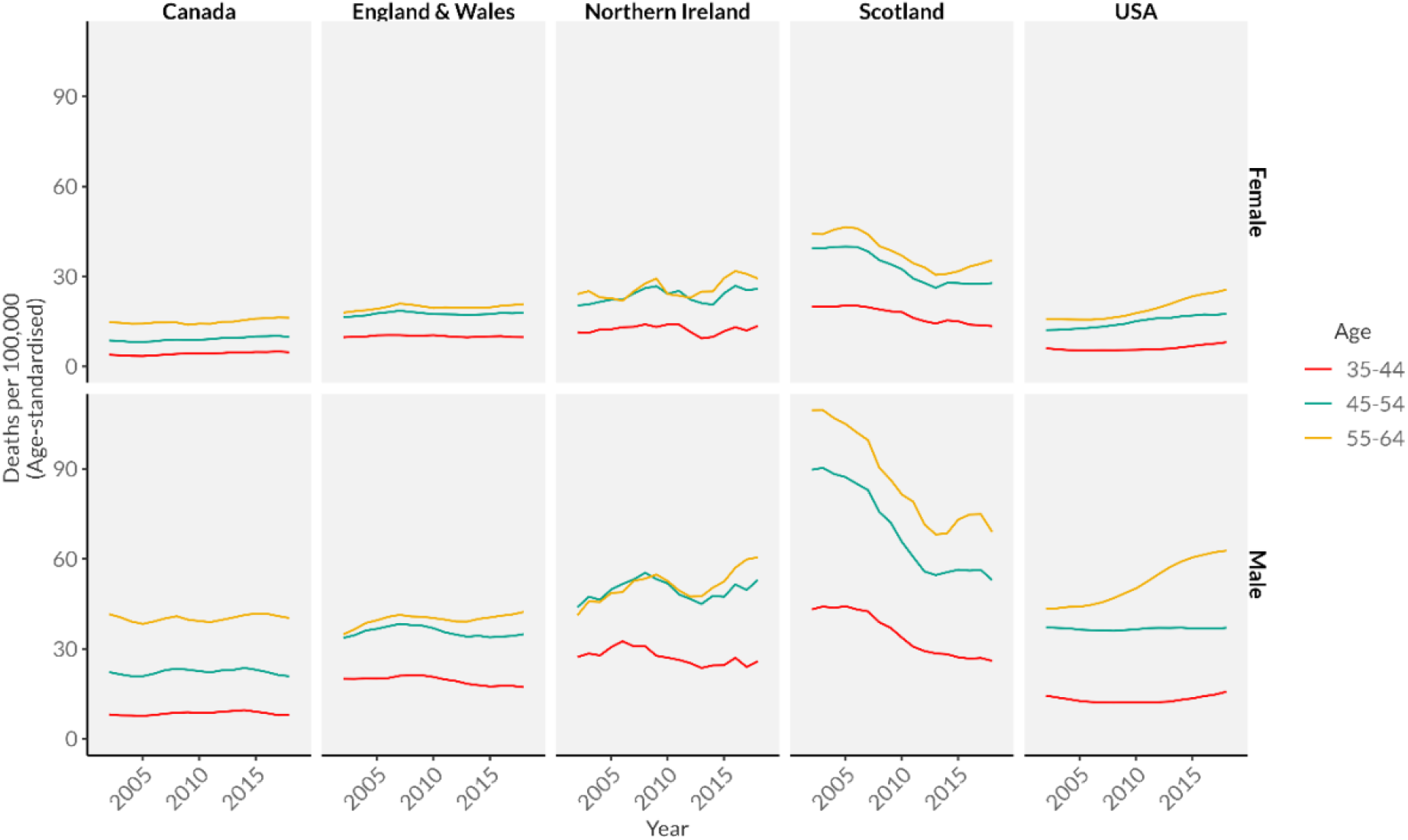
alcohol-specific mortality by age and sex in Canada, the UK and the US, 2001-2019

**Figure S2a.**
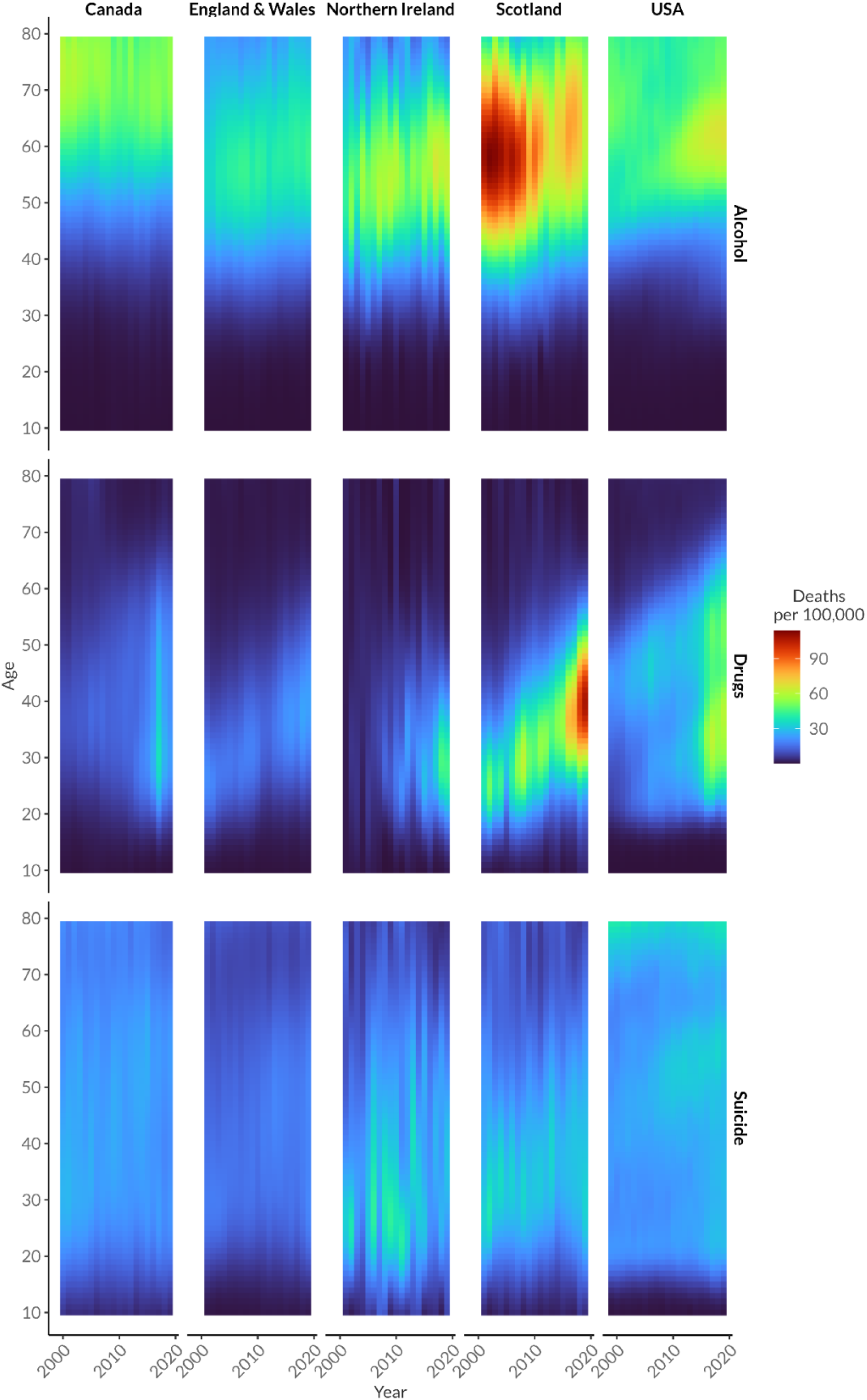
Lexis surface showing age-specific mortality rates for males by cause, year and country

**Figure S2b.**
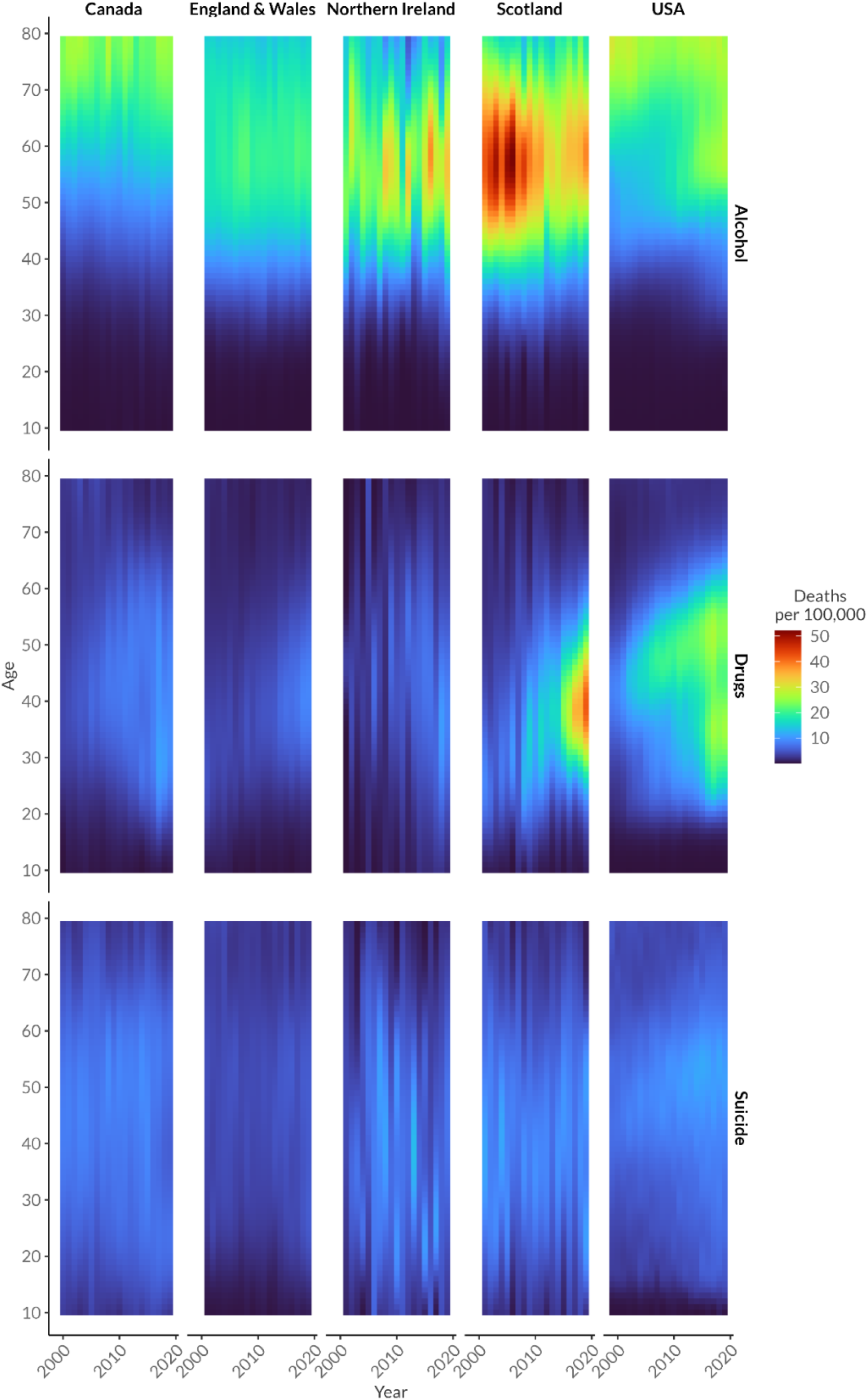
Lexis surface showing age-specific mortality rates for females by cause, year and country

**Table S2.** ICD-10 codes used to define ‘deaths of despair’ categories

| Cause group | ICD-10 codes included |
| --- | --- |
| Suicide | U03, X60-84, Y87 |
| Alcohol-related | K70, K73-74, F10, X45, Y15 |
| Drug-related | F11-16, F18-19, X40-44, X85, Y10-14 |

**Table S2a.** Male mortality rates (per 100,000 population) by cause, age group and year for Canada

| Age group | 2001 | 2002 | 2003 | 2004 | 2005 | 2006 | 2007 | 2008 | 2009 | 2010 | 2011 | 2012 | 2013 | 2014 | 2015 | 2016 | 2017 | 2018 | 2019 |
| --- | --- | --- | --- | --- | --- | --- | --- | --- | --- | --- | --- | --- | --- | --- | --- | --- | --- | --- | --- |
| All-Cause |  |  |  |  |  |  |  |  |  |  |  |  |  |  |  |  |  |  |  |
| 35-44 | 183.6 | 179.4 | 182.6 | 177.9 | 179.1 | 174.9 | 173.3 | 170.6 | 169.0 | 164.1 | 157.8 | 157.5 | 156.2 | 157.2 | 158.0 | 162.4 | 167.9 | 167.9 | 162.0 |
| 45-54 | 424.3 | 419.3 | 415.8 | 404.1 | 399.2 | 388.9 | 388.2 | 383.5 | 373.5 | 361.3 | 350.2 | 347.6 | 347.7 | 344.4 | 340.3 | 344.6 | 342.0 | 344.2 | 334.8 |
| 55-64 | 1156.0 | 1135.9 | 1104.2 | 1075.0 | 1045.1 | 1006.9 | 1010.8 | 988.8 | 949.3 | 917.0 | 901.3 | 886.8 | 884.5 | 877.6 | 860.8 | 863.1 | 845.6 | 845.1 | 825.7 |
| Combined 'Deaths of Despair' |  |  |  |  |  |  |  |  |  |  |  |  |  |  |  |  |  |  |  |
| 35-44 | 44.4 | 43.8 | 44.3 | 42.3 | 47.4 | 43.2 | 44.9 | 45.5 | 47.2 | 46.3 | 45.5 | 45.6 | 47.3 | 49.6 | 51.1 | 51.6 | 61.4 | 51.3 | 50.1 |
| 45-54 | 53.7 | 54.5 | 54.8 | 51.1 | 53.6 | 54.3 | 55.4 | 57.1 | 59.4 | 58.2 | 57.8 | 59.0 | 62.1 | 61.6 | 64.3 | 63.2 | 70.0 | 62.3 | 62.6 |
| 55-64 | 67.9 | 69.9 | 68.6 | 65.0 | 64.0 | 65.4 | 68.5 | 68.0 | 69.8 | 68.3 | 69.0 | 70.5 | 73.6 | 74.2 | 76.8 | 75.2 | 80.0 | 73.3 | 75.5 |
| Alcohol |  |  |  |  |  |  |  |  |  |  |  |  |  |  |  |  |  |  |  |
| 35-44 | 8.2 | 8.0 | 8.2 | 7.2 | 7.9 | 7.8 | 8.2 | 9.1 | 8.9 | 8.5 | 8.6 | 9.0 | 9.6 | 9.2 | 9.8 | 8.2 | 7.7 | 7.9 | 8.4 |
| 45-54 | 22.7 | 22.2 | 21.8 | 20.5 | 20.5 | 21.7 | 23.0 | 24.0 | 23.0 | 22.1 | 22.6 | 21.7 | 24.2 | 23.0 | 23.8 | 22.3 | 20.7 | 20.9 | 21.0 |
| 55-64 | 42.5 | 42.3 | 39.9 | 39.5 | 37.9 | 37.6 | 41.9 | 41.0 | 39.7 | 38.5 | 39.6 | 38.7 | 40.7 | 41.9 | 41.2 | 42.2 | 41.7 | 38.9 | 40.2 |
| Drugs |  |  |  |  |  |  |  |  |  |  |  |  |  |  |  |  |  |  |  |
| 35-44 | 9.7 | 10.5 | 11.0 | 11.2 | 13.4 | 12.5 | 13.7 | 12.4 | 12.9 | 13.4 | 14.3 | 14.0 | 13.8 | 16.0 | 18.3 | 21.8 | 32.1 | 24.0 | 21.1 |
| 45-54 | 6.1 | 7.7 | 7.7 | 8.5 | 9.0 | 9.2 | 10.3 | 10.3 | 11.0 | 11.7 | 12.5 | 13.4 | 12.9 | 13.3 | 15.4 | 18.5 | 26.8 | 20.7 | 19.6 |
| 55-64 | 3.2 | 4.7 | 4.6 | 5.2 | 5.0 | 5.4 | 6.1 | 6.4 | 7.1 | 7.1 | 7.5 | 8.6 | 9.0 | 8.4 | 9.8 | 11.5 | 16.1 | 13.6 | 13.1 |
| Suicide |  |  |  |  |  |  |  |  |  |  |  |  |  |  |  |  |  |  |  |
| 35-44 | 26.6 | 25.3 | 25.1 | 23.8 | 26.1 | 22.9 | 23.1 | 24.0 | 25.4 | 24.4 | 22.6 | 22.6 | 23.8 | 24.4 | 23.0 | 21.7 | 21.5 | 19.4 | 20.6 |
| 45-54 | 24.9 | 24.5 | 25.4 | 22.1 | 24.1 | 23.4 | 22.1 | 22.8 | 25.4 | 24.4 | 22.6 | 23.9 | 25.0 | 25.3 | 25.2 | 22.3 | 22.4 | 20.7 | 22.0 |
| 55-64 | 22.3 | 22.9 | 24.1 | 20.3 | 21.1 | 22.3 | 20.5 | 20.5 | 23.0 | 22.7 | 22.0 | 23.2 | 23.9 | 23.9 | 25.9 | 21.5 | 22.3 | 20.7 | 22.1 |

**Table S2b.**
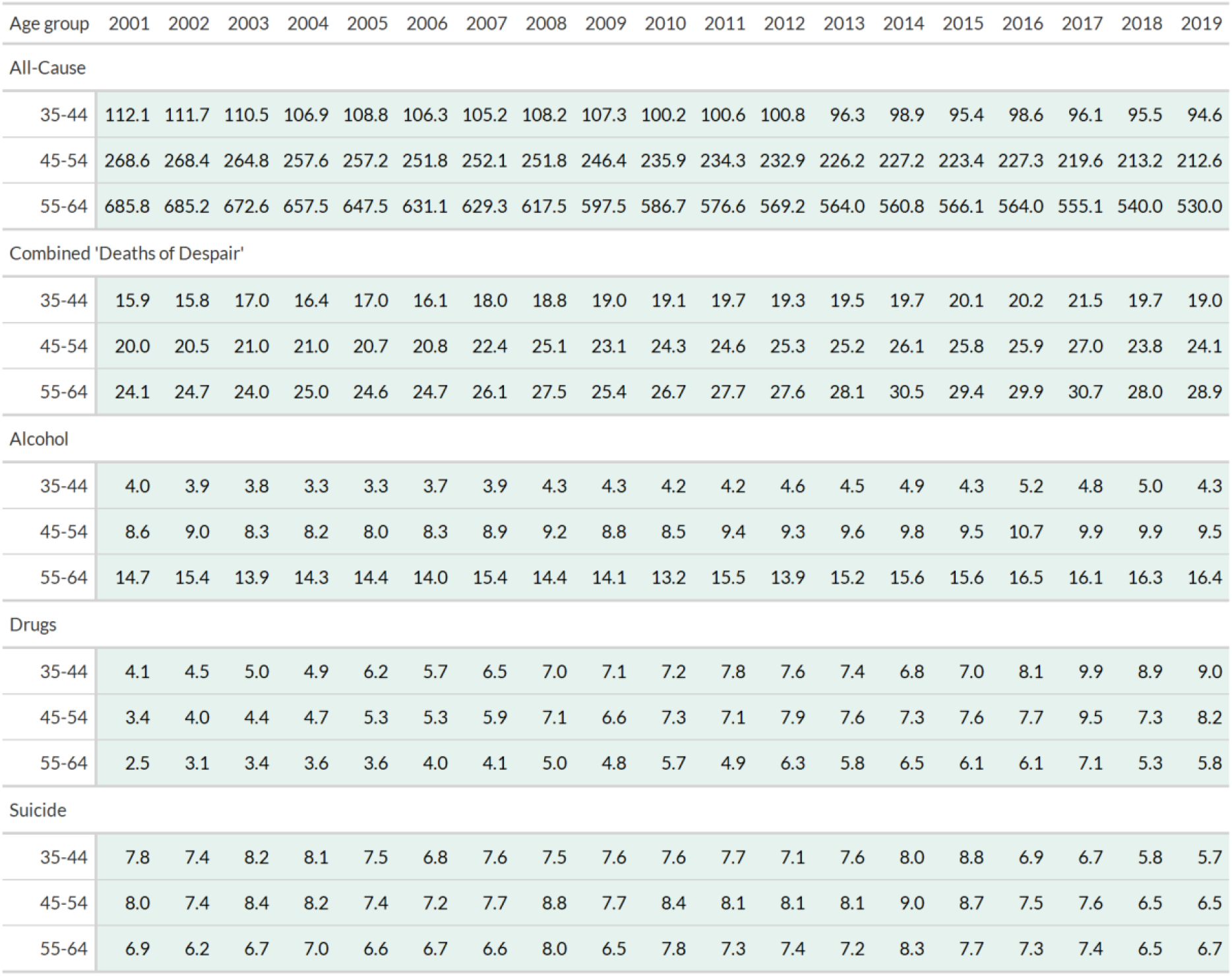
Female mortality rates (per 100,000 population) by cause, age group and year in Canada

| Age group | 2001 | 2002 | 2003 | 2004 | 2005 | 2006 | 2007 | 2008 | 2009 | 2010 | 2011 | 2012 | 2013 | 2014 | 2015 | 2016 | 2017 | 2018 | 2019 |
| --- | --- | --- | --- | --- | --- | --- | --- | --- | --- | --- | --- | --- | --- | --- | --- | --- | --- | --- | --- |
| All-Cause |  |  |  |  |  |  |  |  |  |  |  |  |  |  |  |  |  |  |  |
| 35-44 | 112.1 | 111.7 | 110.5 | 106.9 | 108.8 | 106.3 | 105.2 | 108.2 | 107.3 | 100.2 | 100.6 | 100.8 | 96.3 | 98.9 | 95.4 | 98.6 | 96.1 | 95.5 | 94.6 |
| 45-54 | 268.6 | 268.4 | 264.8 | 257.6 | 257.2 | 251.8 | 252.1 | 251.8 | 246.4 | 235.9 | 234.3 | 232.9 | 226.2 | 227.2 | 223.4 | 227.3 | 219.6 | 213.2 | 212.6 |
| 55-64 | 685.8 | 685.2 | 672.6 | 657.5 | 647.5 | 631.1 | 629.3 | 617.5 | 597.5 | 586.7 | 576.6 | 569.2 | 564.0 | 560.8 | 566.1 | 564.0 | 555.1 | 540.0 | 530.0 |
| Combined 'Deaths of Despair' |  |  |  |  |  |  |  |  |  |  |  |  |  |  |  |  |  |  |  |
| 35-44 | 15.9 | 15.8 | 17.0 | 16.4 | 17.0 | 16.1 | 18.0 | 18.8 | 19.0 | 19.1 | 19.7 | 19.3 | 19.5 | 19.7 | 20.1 | 20.2 | 21.5 | 19.7 | 19.0 |
| 45-54 | 20.0 | 20.5 | 21.0 | 21.0 | 20.7 | 20.8 | 22.4 | 25.1 | 23.1 | 24.3 | 24.6 | 25.3 | 25.2 | 26.1 | 25.8 | 25.9 | 27.0 | 23.8 | 24.1 |
| 55-64 | 24.1 | 24.7 | 24.0 | 25.0 | 24.6 | 24.7 | 26.1 | 27.5 | 25.4 | 26.7 | 27.7 | 27.6 | 28.1 | 30.5 | 29.4 | 29.9 | 30.7 | 28.0 | 28.9 |
| Alcohol |  |  |  |  |  |  |  |  |  |  |  |  |  |  |  |  |  |  |  |
| 35-44 | 4.0 | 3.9 | 3.8 | 3.3 | 3.3 | 3.7 | 3.9 | 4.3 | 4.3 | 4.2 | 4.2 | 4.6 | 4.5 | 4.9 | 4.3 | 5.2 | 4.8 | 5.0 | 4.3 |
| 45-54 | 8.6 | 9.0 | 8.3 | 8.2 | 8.0 | 8.3 | 8.9 | 9.2 | 8.8 | 8.5 | 9.4 | 9.3 | 9.6 | 9.8 | 9.5 | 10.7 | 9.9 | 9.9 | 9.5 |
| 55-64 | 14.7 | 15.4 | 13.9 | 14.3 | 14.4 | 14.0 | 15.4 | 14.4 | 14.1 | 13.2 | 15.5 | 13.9 | 15.2 | 15.6 | 15.6 | 16.5 | 16.1 | 16.3 | 16.4 |
| Drugs |  |  |  |  |  |  |  |  |  |  |  |  |  |  |  |  |  |  |  |
| 35-44 | 4.1 | 4.5 | 5.0 | 4.9 | 6.2 | 5.7 | 6.5 | 7.0 | 7.1 | 7.2 | 7.8 | 7.6 | 7.4 | 6.8 | 7.0 | 8.1 | 9.9 | 8.9 | 9.0 |
| 45-54 | 3.4 | 4.0 | 4.4 | 4.7 | 5.3 | 5.3 | 5.9 | 7.1 | 6.6 | 7.3 | 7.1 | 7.9 | 7.6 | 7.3 | 7.6 | 7.7 | 9.5 | 7.3 | 8.2 |
| 55-64 | 2.5 | 3.1 | 3.4 | 3.6 | 3.6 | 4.0 | 4.1 | 5.0 | 4.8 | 5.7 | 4.9 | 6.3 | 5.8 | 6.5 | 6.1 | 6.1 | 7.1 | 5.3 | 5.8 |
| Suicide |  |  |  |  |  |  |  |  |  |  |  |  |  |  |  |  |  |  |  |
| 35-44 | 7.8 | 7.4 | 8.2 | 8.1 | 7.5 | 6.8 | 7.6 | 7.5 | 7.6 | 7.6 | 7.7 | 7.1 | 7.6 | 8.0 | 8.8 | 6.9 | 6.7 | 5.8 | 5.7 |
| 45-54 | 8.0 | 7.4 | 8.4 | 8.2 | 7.4 | 7.2 | 7.7 | 8.8 | 7.7 | 8.4 | 8.1 | 8.1 | 8.1 | 9.0 | 8.7 | 7.5 | 7.6 | 6.5 | 6.5 |
| 55-64 | 6.9 | 6.2 | 6.7 | 7.0 | 6.6 | 6.7 | 6.6 | 8.0 | 6.5 | 7.8 | 7.3 | 7.4 | 7.2 | 8.3 | 7.7 | 7.3 | 7.4 | 6.5 | 6.7 |

**Table S3a.** Male mortality rates (per 100,000 population) by cause, age group and year for England & Wales

| Age group | 2001 | 2002 | 2003 | 2004 | 2005 | 2006 | 2007 | 2008 | 2009 | 2010 | 2011 | 2012 | 2013 | 2014 | 2015 | 2016 | 2017 | 2018 | 2019 |
| --- | --- | --- | --- | --- | --- | --- | --- | --- | --- | --- | --- | --- | --- | --- | --- | --- | --- | --- | --- |
| All-Cause |  |  |  |  |  |  |  |  |  |  |  |  |  |  |  |  |  |  |  |
| 35-44 | 190.7 | 188.3 | 188.4 | 183.7 | 181.3 | 181.5 | 177.1 | 179.8 | 177.7 | 168.9 | 162.8 | 155.2 | 161.4 | 159.5 | 160.7 | 162.9 | 161.7 | 168.0 | 162.8 |
| 45-54 | 461.4 | 453.4 | 451.1 | 433.0 | 428.0 | 424.2 | 412.7 | 410.8 | 402.8 | 393.9 | 382.2 | 368.3 | 375.6 | 370.5 | 372.9 | 374.7 | 374.4 | 382.0 | 371.0 |
| 55-64 | 1280.1 | 1251.8 | 1226.2 | 1159.3 | 1129.0 | 1100.6 | 1070.2 | 1044.7 | 1003.7 | 991.9 | 954.9 | 929.1 | 926.8 | 913.1 | 917.4 | 916.9 | 907.3 | 910.9 | 888.6 |
| Combined 'Deaths of Despair' |  |  |  |  |  |  |  |  |  |  |  |  |  |  |  |  |  |  |  |
| 35-44 | 44.8 | 42.8 | 44.4 | 44.7 | 45.4 | 46.7 | 46.2 | 51.0 | 51.5 | 48.0 | 49.2 | 47.9 | 51.2 | 52.1 | 54.1 | 54.5 | 54.1 | 58.3 | 58.0 |
| 45-54 | 50.5 | 48.7 | 52.0 | 53.5 | 55.0 | 56.6 | 56.6 | 59.6 | 58.4 | 58.0 | 59.0 | 55.9 | 61.5 | 62.0 | 63.3 | 62.0 | 64.5 | 66.4 | 71.1 |
| 55-64 | 46.6 | 45.9 | 49.5 | 51.3 | 52.8 | 53.6 | 54.4 | 56.1 | 53.1 | 56.0 | 56.3 | 52.5 | 57.6 | 59.3 | 59.6 | 58.9 | 60.9 | 61.6 | 66.1 |
| Alcohol |  |  |  |  |  |  |  |  |  |  |  |  |  |  |  |  |  |  |  |
| 35-44 | 19.8 | 19.1 | 21.0 | 19.7 | 19.5 | 21.0 | 20.3 | 21.9 | 21.4 | 20.3 | 20.2 | 18.9 | 18.7 | 17.4 | 17.5 | 17.5 | 17.9 | 17.5 | 16.3 |
| 45-54 | 32.7 | 32.0 | 36.1 | 35.5 | 36.7 | 37.9 | 38.0 | 39.0 | 36.9 | 37.4 | 36.4 | 32.8 | 35.0 | 34.3 | 33.7 | 33.5 | 35.1 | 34.3 | 35.3 |
| 55-64 | 33.1 | 33.8 | 37.5 | 38.2 | 40.1 | 40.3 | 41.6 | 42.1 | 38.7 | 41.4 | 40.7 | 37.2 | 39.4 | 40.6 | 40.0 | 40.8 | 42.3 | 41.3 | 43.3 |
| Drugs |  |  |  |  |  |  |  |  |  |  |  |  |  |  |  |  |  |  |  |
| 35-44 | 9.8 | 8.2 | 8.6 | 9.5 | 10.4 | 10.1 | 11.3 | 13.6 | 14.0 | 12.8 | 11.8 | 12.1 | 14.4 | 16.9 | 19.9 | 20.6 | 19.9 | 23.3 | 22.8 |
| 45-54 | 4.6 | 3.7 | 3.6 | 4.3 | 4.8 | 4.5 | 5.6 | 6.4 | 6.7 | 6.1 | 6.3 | 6.6 | 7.8 | 9.7 | 12.1 | 11.8 | 12.9 | 15.0 | 16.6 |
| 55-64 | 2.3 | 1.8 | 1.6 | 1.9 | 2.2 | 1.9 | 2.3 | 2.4 | 2.6 | 2.5 | 2.8 | 2.6 | 3.1 | 3.9 | 4.8 | 4.3 | 5.0 | 6.0 | 6.5 |
| Suicide |  |  |  |  |  |  |  |  |  |  |  |  |  |  |  |  |  |  |  |
| 35-44 | 15.2 | 15.4 | 14.8 | 15.5 | 15.4 | 15.6 | 14.7 | 15.5 | 16.1 | 14.9 | 17.2 | 16.9 | 18.1 | 17.8 | 16.7 | 16.4 | 16.3 | 17.4 | 18.9 |
| 45-54 | 13.2 | 13.0 | 12.3 | 13.7 | 13.5 | 14.2 | 12.9 | 14.2 | 14.8 | 14.5 | 16.2 | 16.5 | 18.7 | 17.9 | 17.5 | 16.7 | 16.5 | 17.1 | 19.1 |
| 55-64 | 11.1 | 10.3 | 10.3 | 11.2 | 10.5 | 11.4 | 10.5 | 11.6 | 11.8 | 12.1 | 12.8 | 12.8 | 15.1 | 14.7 | 14.8 | 13.8 | 13.5 | 14.3 | 16.3 |

**Table S3b.** Female mortality rates (per 100,000 population) by cause, age group and year in England & Wales

| Age group | 2001 | 2002 | 2003 | 2004 | 2005 | 2006 | 2007 | 2008 | 2009 | 2010 | 2011 | 2012 | 2013 | 2014 | 2015 | 2016 | 2017 | 2018 | 2019 |
| --- | --- | --- | --- | --- | --- | --- | --- | --- | --- | --- | --- | --- | --- | --- | --- | --- | --- | --- | --- |
| All-Cause |  |  |  |  |  |  |  |  |  |  |  |  |  |  |  |  |  |  |  |
| 35-44 | 117.5 | 115.0 | 116.4 | 112.2 | 111.6 | 110.4 | 109.3 | 110.2 | 105.6 | 104.8 | 100.9 | 97.4 | 97.8 | 98.8 | 99.0 | 99.3 | 97.8 | 99.2 | 98.7 |
| 45-54 | 302.5 | 297.0 | 295.1 | 282.4 | 281.3 | 276.6 | 273.2 | 272.8 | 262.3 | 260.4 | 253.5 | 249.5 | 246.7 | 244.2 | 247.4 | 251.2 | 245.3 | 248.1 | 243.0 |
| 55-64 | 802.5 | 784.8 | 770.3 | 735.2 | 722.1 | 700.6 | 687.0 | 680.8 | 651.2 | 641.2 | 629.7 | 623.5 | 612.7 | 600.7 | 608.3 | 621.5 | 599.8 | 608.3 | 587.8 |
| Combined 'Deaths of Despair' |  |  |  |  |  |  |  |  |  |  |  |  |  |  |  |  |  |  |  |
| 35-44 | 16.8 | 16.7 | 17.1 | 17.7 | 18.2 | 17.5 | 17.3 | 18.7 | 17.7 | 18.3 | 19.1 | 18.0 | 18.8 | 20.7 | 21.4 | 20.4 | 21.6 | 22.2 | 23.2 |
| 45-54 | 23.0 | 22.6 | 23.0 | 24.1 | 24.7 | 25.5 | 24.7 | 26.0 | 24.5 | 24.6 | 25.5 | 24.9 | 25.3 | 26.9 | 27.7 | 28.2 | 28.5 | 28.9 | 30.5 |
| 55-64 | 23.7 | 23.0 | 24.0 | 25.1 | 24.3 | 26.3 | 26.3 | 26.9 | 24.9 | 25.4 | 25.9 | 25.6 | 25.4 | 26.8 | 27.4 | 27.4 | 28.6 | 28.8 | 29.4 |
| Alcohol |  |  |  |  |  |  |  |  |  |  |  |  |  |  |  |  |  |  |  |
| 35-44 | 9.6 | 9.8 | 9.6 | 10.0 | 10.2 | 10.8 | 10.2 | 10.2 | 10.4 | 10.2 | 10.6 | 9.5 | 9.4 | 10.1 | 10.1 | 9.8 | 10.4 | 9.1 | 9.7 |
| 45-54 | 16.4 | 16.4 | 16.4 | 17.3 | 17.3 | 18.5 | 18.4 | 18.8 | 17.3 | 17.3 | 17.6 | 17.2 | 17.2 | 17.2 | 17.5 | 17.5 | 18.5 | 17.2 | 18.2 |
| 55-64 | 17.7 | 17.4 | 18.7 | 19.1 | 18.4 | 20.1 | 21.2 | 21.5 | 19.0 | 19.6 | 19.7 | 19.7 | 19.4 | 19.6 | 19.9 | 19.6 | 21.1 | 20.6 | 20.5 |
| Drugs |  |  |  |  |  |  |  |  |  |  |  |  |  |  |  |  |  |  |  |
| 35-44 | 3.5 | 3.4 | 3.5 | 3.4 | 3.8 | 2.9 | 3.4 | 4.4 | 3.7 | 4.3 | 4.6 | 4.7 | 4.9 | 6.2 | 6.7 | 6.6 | 7.1 | 8.3 | 8.4 |
| 45-54 | 2.5 | 2.5 | 2.6 | 2.5 | 2.9 | 2.4 | 2.5 | 3.1 | 3.2 | 3.3 | 3.5 | 3.6 | 3.7 | 4.8 | 5.0 | 5.7 | 5.8 | 6.6 | 7.1 |
| 55-64 | 1.9 | 1.8 | 1.8 | 1.8 | 1.9 | 1.9 | 1.6 | 1.8 | 2.0 | 2.1 | 2.2 | 2.2 | 2.3 | 2.9 | 2.8 | 3.2 | 3.4 | 3.5 | 4.2 |
| Suicide |  |  |  |  |  |  |  |  |  |  |  |  |  |  |  |  |  |  |  |
| 35-44 | 3.8 | 3.5 | 4.0 | 4.2 | 4.2 | 3.8 | 3.6 | 4.2 | 3.7 | 3.8 | 3.9 | 3.9 | 4.5 | 4.4 | 4.6 | 4.1 | 4.1 | 4.8 | 5.2 |
| 45-54 | 4.2 | 3.7 | 4.0 | 4.3 | 4.5 | 4.5 | 3.8 | 4.1 | 4.0 | 4.0 | 4.4 | 4.1 | 4.4 | 4.9 | 5.1 | 5.1 | 4.3 | 5.2 | 5.3 |
| 55-64 | 4.1 | 3.8 | 3.6 | 4.1 | 4.0 | 4.3 | 3.6 | 3.6 | 3.8 | 3.7 | 3.9 | 3.7 | 3.6 | 4.4 | 4.8 | 4.6 | 4.0 | 4.6 | 4.7 |

**Table S4a.** Male mortality rates (per 100,000 population) by cause, age group and year for Northern Ireland

| Age group | 2001 | 2002 | 2003 | 2004 | 2005 | 2006 | 2007 | 2008 | 2009 | 2010 | 2011 | 2012 | 2013 | 2014 | 2015 | 2016 | 2017 | 2018 | 2019 |
| --- | --- | --- | --- | --- | --- | --- | --- | --- | --- | --- | --- | --- | --- | --- | --- | --- | --- | --- | --- |
| All-Cause |  |  |  |  |  |  |  |  |  |  |  |  |  |  |  |  |  |  |  |
| 35-44 | 192.7 | 200.4 | 170.8 | 199.3 | 199.3 | 211.6 | 223.7 | 199.2 | 204.4 | 207.7 | 177.2 | 189.6 | 188.9 | 173.4 | 184.3 | 172.7 | 193.7 | 183.7 | 190.4 |
| 45-54 | 465.7 | 497.4 | 438.8 | 479.3 | 450.5 | 475.5 | 480.3 | 440.9 | 431.9 | 443.7 | 396.4 | 398.5 | 417.9 | 375.3 | 393.1 | 388.9 | 405.0 | 395.8 | 378.1 |
| 55-64 | 1324.3 | 1385.5 | 1282.8 | 1282.1 | 1214.4 | 1205.1 | 1170.8 | 1137.3 | 1067.7 | 1092.9 | 1042.6 | 971.1 | 1011.6 | 940.0 | 977.1 | 977.9 | 967.5 | 947.7 | 876.9 |
| Combined 'Deaths of Despair' |  |  |  |  |  |  |  |  |  |  |  |  |  |  |  |  |  |  |  |
| 35-44 | 47.6 | 60.2 | 45.0 | 54.7 | 63.8 | 71.6 | 71.1 | 71.1 | 67.5 | 67.9 | 56.7 | 68.4 | 74.4 | 62.6 | 70.0 | 59.9 | 74.7 | 70.4 | 81.9 |
| 45-54 | 54.9 | 70.2 | 57.4 | 70.5 | 73.3 | 83.4 | 86.3 | 83.5 | 85.9 | 82.6 | 73.9 | 74.7 | 86.3 | 68.5 | 88.3 | 71.5 | 86.9 | 81.0 | 85.0 |
| 55-64 | 50.0 | 57.0 | 55.2 | 64.2 | 62.5 | 73.8 | 74.8 | 76.1 | 72.6 | 79.6 | 67.9 | 65.3 | 77.3 | 64.1 | 79.3 | 73.3 | 79.6 | 83.7 | 74.5 |
| Alcohol |  |  |  |  |  |  |  |  |  |  |  |  |  |  |  |  |  |  |  |
| 35-44 | 26.7 | 34.7 | 20.5 | 30.1 | 32.8 | 28.9 | 35.9 | 27.6 | 29.1 | 26.3 | 25.8 | 26.8 | 23.1 | 21.0 | 29.4 | 23.3 | 28.4 | 20.2 | 29.0 |
| 45-54 | 41.2 | 51.3 | 39.1 | 51.7 | 48.3 | 49.5 | 57.1 | 52.9 | 56.2 | 50.8 | 48.5 | 45.2 | 46.3 | 43.4 | 53.3 | 45.4 | 55.8 | 47.7 | 55.5 |
| 55-64 | 38.1 | 44.0 | 41.5 | 52.0 | 43.5 | 50.5 | 52.7 | 54.7 | 52.8 | 56.8 | 48.2 | 43.4 | 50.6 | 48.5 | 52.1 | 56.7 | 62.3 | 60.5 | 58.8 |
| Drugs |  |  |  |  |  |  |  |  |  |  |  |  |  |  |  |  |  |  |  |
| 35-44 | 1.6 | 4.1 | 4.1 | 3.7 | 4.6 | 6.5 | 7.2 | 9.2 | 7.3 | 10.6 | 9.4 | 18.0 | 15.3 | 13.6 | 16.1 | 16.5 | 19.6 | 26.9 | 23.6 |
| 45-54 | 1.8 | 2.2 | 3.2 | 3.8 | 3.5 | 4.2 | 3.8 | 5.1 | 4.7 | 6.3 | 5.8 | 6.3 | 7.4 | 5.0 | 9.7 | 8.3 | 7.5 | 9.4 | 6.2 |
| 55-64 | 2.0 | 1.2 | 2.5 | 2.6 | 2.4 | 2.0 | 2.0 | 2.6 | 1.0 | 4.1 | 1.5 | 1.2 | 2.6 | 1.7 | 4.6 | 2.8 | 3.0 | 3.4 | 0.8 |
| Suicide |  |  |  |  |  |  |  |  |  |  |  |  |  |  |  |  |  |  |  |
| 35-44 | 19.4 | 21.4 | 20.4 | 20.8 | 26.4 | 36.2 | 27.9 | 34.2 | 31.1 | 31.0 | 21.4 | 23.5 | 36.0 | 28.1 | 24.5 | 20.1 | 26.6 | 23.2 | 29.3 |
| 45-54 | 11.9 | 16.7 | 15.0 | 15.0 | 21.5 | 29.7 | 25.3 | 25.5 | 25.0 | 25.5 | 19.6 | 23.3 | 32.6 | 20.1 | 25.3 | 17.7 | 23.6 | 24.0 | 23.4 |
| 55-64 | 9.9 | 11.8 | 11.2 | 9.6 | 16.6 | 21.3 | 20.1 | 18.7 | 18.7 | 18.7 | 18.2 | 20.6 | 24.1 | 13.8 | 22.6 | 13.8 | 14.3 | 19.9 | 15.0 |

**Table S4b.**
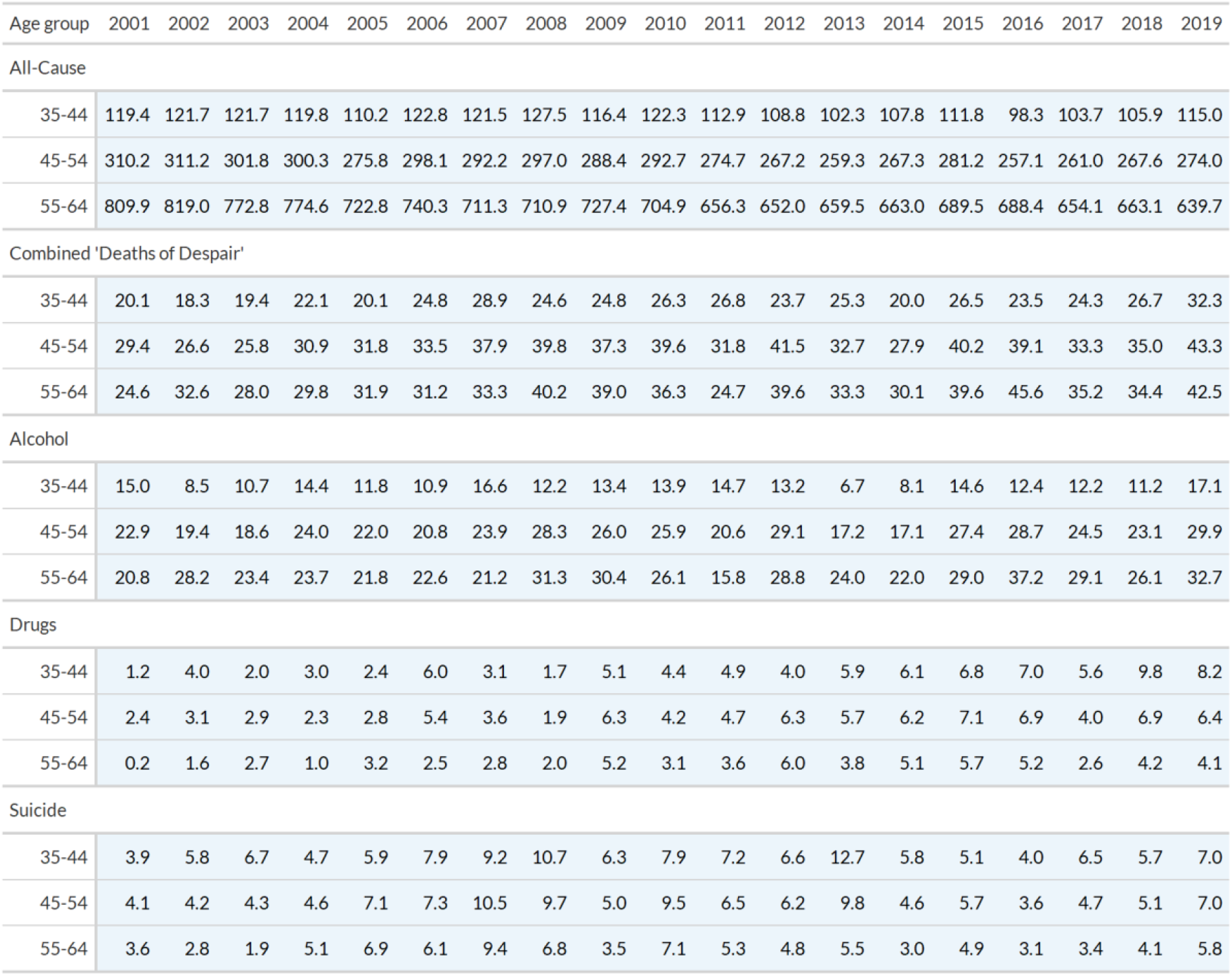
Female mortality rates (per 100,000 population) by cause, age group and year in Northern Ireland

| Age group | 2001 | 2002 | 2003 | 2004 | 2005 | 2006 | 2007 | 2008 | 2009 | 2010 | 2011 | 2012 | 2013 | 2014 | 2015 | 2016 | 2017 | 2018 | 2019 |
| --- | --- | --- | --- | --- | --- | --- | --- | --- | --- | --- | --- | --- | --- | --- | --- | --- | --- | --- | --- |
| All-Cause |  |  |  |  |  |  |  |  |  |  |  |  |  |  |  |  |  |  |  |
| 35-44 | 119.4 | 121.7 | 121.7 | 119.8 | 110.2 | 122.8 | 121.5 | 127.5 | 116.4 | 122.3 | 112.9 | 108.8 | 102.3 | 107.8 | 111.8 | 98.3 | 103.7 | 105.9 | 115.0 |
| 45-54 | 310.2 | 311.2 | 301.8 | 300.3 | 275.8 | 298.1 | 292.2 | 297.0 | 288.4 | 292.7 | 274.7 | 267.2 | 259.3 | 267.3 | 281.2 | 257.1 | 261.0 | 267.6 | 274.0 |
| 55-64 | 809.9 | 819.0 | 772.8 | 774.6 | 722.8 | 740.3 | 711.3 | 710.9 | 727.4 | 704.9 | 656.3 | 652.0 | 659.5 | 663.0 | 689.5 | 688.4 | 654.1 | 663.1 | 639.7 |
| Combined 'Deaths of Despair' |  |  |  |  |  |  |  |  |  |  |  |  |  |  |  |  |  |  |  |
| 35-44 | 20.1 | 18.3 | 19.4 | 22.1 | 20.1 | 24.8 | 28.9 | 24.6 | 24.8 | 26.3 | 26.8 | 23.7 | 25.3 | 20.0 | 26.5 | 23.5 | 24.3 | 26.7 | 32.3 |
| 45-54 | 29.4 | 26.6 | 25.8 | 30.9 | 31.8 | 33.5 | 37.9 | 39.8 | 37.3 | 39.6 | 31.8 | 41.5 | 32.7 | 27.9 | 40.2 | 39.1 | 33.3 | 35.0 | 43.3 |
| 55-64 | 24.6 | 32.6 | 28.0 | 29.8 | 31.9 | 31.2 | 33.3 | 40.2 | 39.0 | 36.3 | 24.7 | 39.6 | 33.3 | 30.1 | 39.6 | 45.6 | 35.2 | 34.4 | 42.5 |
| Alcohol |  |  |  |  |  |  |  |  |  |  |  |  |  |  |  |  |  |  |  |
| 35-44 | 15.0 | 8.5 | 10.7 | 14.4 | 11.8 | 10.9 | 16.6 | 12.2 | 13.4 | 13.9 | 14.7 | 13.2 | 6.7 | 8.1 | 14.6 | 12.4 | 12.2 | 11.2 | 17.1 |
| 45-54 | 22.9 | 19.4 | 18.6 | 24.0 | 22.0 | 20.8 | 23.9 | 28.3 | 26.0 | 25.9 | 20.6 | 29.1 | 17.2 | 17.1 | 27.4 | 28.7 | 24.5 | 23.1 | 29.9 |
| 55-64 | 20.8 | 28.2 | 23.4 | 23.7 | 21.8 | 22.6 | 21.2 | 31.3 | 30.4 | 26.1 | 15.8 | 28.8 | 24.0 | 22.0 | 29.0 | 37.2 | 29.1 | 26.1 | 32.7 |
| Drugs |  |  |  |  |  |  |  |  |  |  |  |  |  |  |  |  |  |  |  |
| 35-44 | 1.2 | 4.0 | 2.0 | 3.0 | 2.4 | 6.0 | 3.1 | 1.7 | 5.1 | 4.4 | 4.9 | 4.0 | 5.9 | 6.1 | 6.8 | 7.0 | 5.6 | 9.8 | 8.2 |
| 45-54 | 2.4 | 3.1 | 2.9 | 2.3 | 2.8 | 5.4 | 3.6 | 1.9 | 6.3 | 4.2 | 4.7 | 6.3 | 5.7 | 6.2 | 7.1 | 6.9 | 4.0 | 6.9 | 6.4 |
| 55-64 | 0.2 | 1.6 | 2.7 | 1.0 | 3.2 | 2.5 | 2.8 | 2.0 | 5.2 | 3.1 | 3.6 | 6.0 | 3.8 | 5.1 | 5.7 | 5.2 | 2.6 | 4.2 | 4.1 |
| Suicide |  |  |  |  |  |  |  |  |  |  |  |  |  |  |  |  |  |  |  |
| 35-44 | 3.9 | 5.8 | 6.7 | 4.7 | 5.9 | 7.9 | 9.2 | 10.7 | 6.3 | 7.9 | 7.2 | 6.6 | 12.7 | 5.8 | 5.1 | 4.0 | 6.5 | 5.7 | 7.0 |
| 45-54 | 4.1 | 4.2 | 4.3 | 4.6 | 7.1 | 7.3 | 10.5 | 9.7 | 5.0 | 9.5 | 6.5 | 6.2 | 9.8 | 4.6 | 5.7 | 3.6 | 4.7 | 5.1 | 7.0 |
| 55-64 | 3.6 | 2.8 | 1.9 | 5.1 | 6.9 | 6.1 | 9.4 | 6.8 | 3.5 | 7.1 | 5.3 | 4.8 | 5.5 | 3.0 | 4.9 | 3.1 | 3.4 | 4.1 | 5.8 |

**Table S5a.**
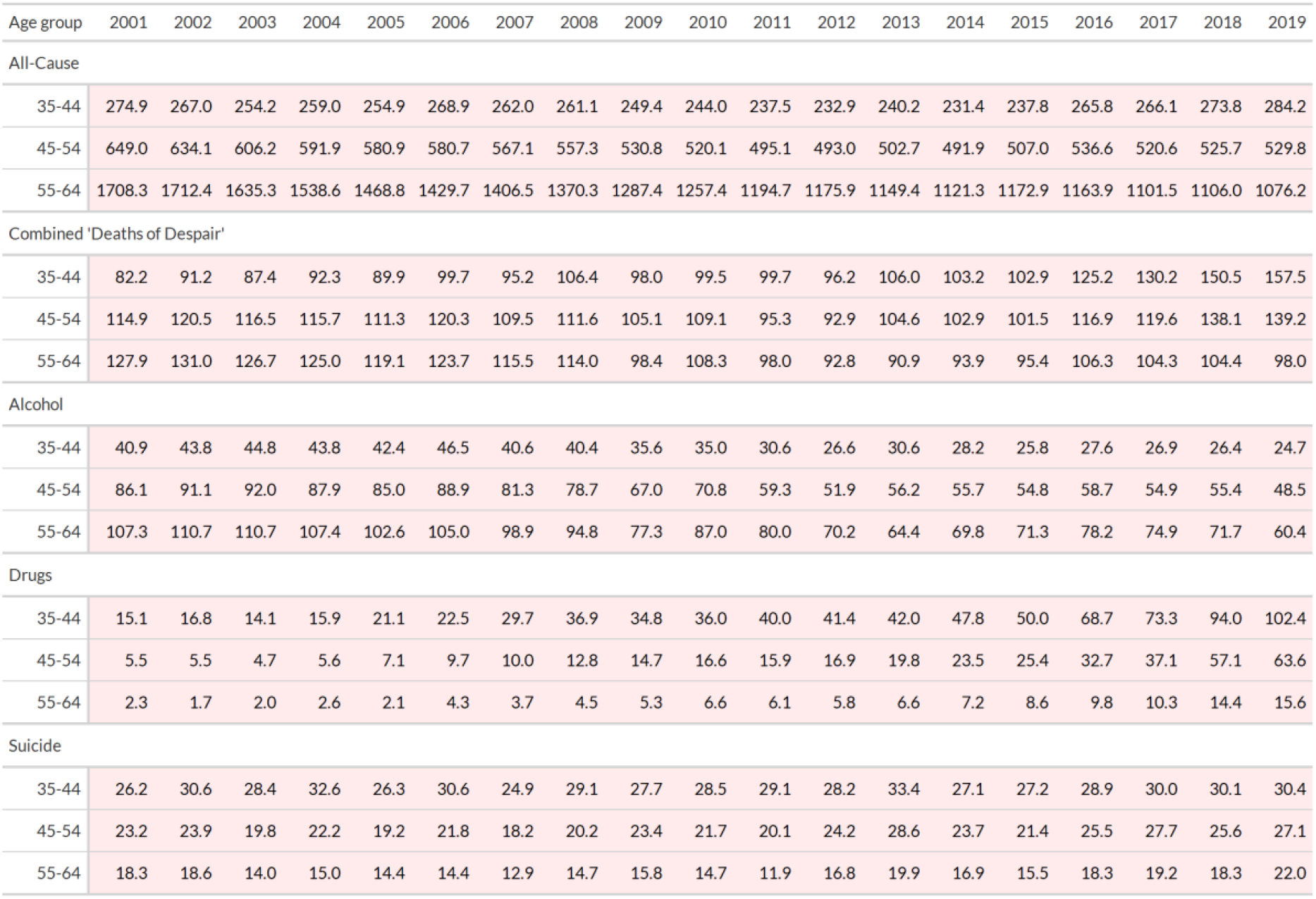
Male mortality rates (per 100,000 population) by cause, age group and year for Scotland

| Age group | 2001 | 2002 | 2003 | 2004 | 2005 | 2006 | 2007 | 2008 | 2009 | 2010 | 2011 | 2012 | 2013 | 2014 | 2015 | 2016 | 2017 | 2018 | 2019 |
| --- | --- | --- | --- | --- | --- | --- | --- | --- | --- | --- | --- | --- | --- | --- | --- | --- | --- | --- | --- |
| All-Cause |  |  |  |  |  |  |  |  |  |  |  |  |  |  |  |  |  |  |  |
| 35-44 | 274.9 | 267.0 | 254.2 | 259.0 | 254.9 | 268.9 | 262.0 | 261.1 | 249.4 | 244.0 | 237.5 | 232.9 | 240.2 | 231.4 | 237.8 | 265.8 | 266.1 | 273.8 | 284.2 |
| 45-54 | 649.0 | 634.1 | 606.2 | 591.9 | 580.9 | 580.7 | 567.1 | 557.3 | 530.8 | 520.1 | 495.1 | 493.0 | 502.7 | 491.9 | 507.0 | 536.6 | 520.6 | 525.7 | 529.8 |
| 55-64 | 1708.3 | 1712.4 | 1635.3 | 1538.6 | 1468.8 | 1429.7 | 1406.5 | 1370.3 | 1287.4 | 1257.4 | 1194.7 | 1175.9 | 1149.4 | 1121.3 | 1172.9 | 1163.9 | 1101.5 | 1106.0 | 1076.2 |
| Combined 'Deaths of Despair' |  |  |  |  |  |  |  |  |  |  |  |  |  |  |  |  |  |  |  |
| 35-44 | 82.2 | 91.2 | 87.4 | 92.3 | 89.9 | 99.7 | 95.2 | 106.4 | 98.0 | 99.5 | 99.7 | 96.2 | 106.0 | 103.2 | 102.9 | 125.2 | 130.2 | 150.5 | 157.5 |
| 45-54 | 114.9 | 120.5 | 116.5 | 115.7 | 111.3 | 120.3 | 109.5 | 111.6 | 105.1 | 109.1 | 95.3 | 92.9 | 104.6 | 102.9 | 101.5 | 116.9 | 119.6 | 138.1 | 139.2 |
| 55-64 | 127.9 | 131.0 | 126.7 | 125.0 | 119.1 | 123.7 | 115.5 | 114.0 | 98.4 | 108.3 | 98.0 | 92.8 | 90.9 | 93.9 | 95.4 | 106.3 | 104.3 | 104.4 | 98.0 |
| Alcohol |  |  |  |  |  |  |  |  |  |  |  |  |  |  |  |  |  |  |  |
| 35-44 | 40.9 | 43.8 | 44.8 | 43.8 | 42.4 | 46.5 | 40.6 | 40.4 | 35.6 | 35.0 | 30.6 | 26.6 | 30.6 | 28.2 | 25.8 | 27.6 | 26.9 | 26.4 | 24.7 |
| 45-54 | 86.1 | 91.1 | 92.0 | 87.9 | 85.0 | 88.9 | 81.3 | 78.7 | 67.0 | 70.8 | 59.3 | 51.9 | 56.2 | 55.7 | 54.8 | 58.7 | 54.9 | 55.4 | 48.5 |
| 55-64 | 107.3 | 110.7 | 110.7 | 107.4 | 102.6 | 105.0 | 98.9 | 94.8 | 77.3 | 87.0 | 80.0 | 70.2 | 64.4 | 69.8 | 71.3 | 78.2 | 74.9 | 71.7 | 60.4 |
| Drugs |  |  |  |  |  |  |  |  |  |  |  |  |  |  |  |  |  |  |  |
| 35-44 | 15.1 | 16.8 | 14.1 | 15.9 | 21.1 | 22.5 | 29.7 | 36.9 | 34.8 | 36.0 | 40.0 | 41.4 | 42.0 | 47.8 | 50.0 | 68.7 | 73.3 | 94.0 | 102.4 |
| 45-54 | 5.5 | 5.5 | 4.7 | 5.6 | 7.1 | 9.7 | 10.0 | 12.8 | 14.7 | 16.6 | 15.9 | 16.9 | 19.8 | 23.5 | 25.4 | 32.7 | 37.1 | 57.1 | 63.6 |
| 55-64 | 2.3 | 1.7 | 2.0 | 2.6 | 2.1 | 4.3 | 3.7 | 4.5 | 5.3 | 6.6 | 6.1 | 5.8 | 6.6 | 7.2 | 8.6 | 9.8 | 10.3 | 14.4 | 15.6 |
| Suicide |  |  |  |  |  |  |  |  |  |  |  |  |  |  |  |  |  |  |  |
| 35-44 | 26.2 | 30.6 | 28.4 | 32.6 | 26.3 | 30.6 | 24.9 | 29.1 | 27.7 | 28.5 | 29.1 | 28.2 | 33.4 | 27.1 | 27.2 | 28.9 | 30.0 | 30.1 | 30.4 |
| 45-54 | 23.2 | 23.9 | 19.8 | 22.2 | 19.2 | 21.8 | 18.2 | 20.2 | 23.4 | 21.7 | 20.1 | 24.2 | 28.6 | 23.7 | 21.4 | 25.5 | 27.7 | 25.6 | 27.1 |
| 55-64 | 18.3 | 18.6 | 14.0 | 15.0 | 14.4 | 14.4 | 12.9 | 14.7 | 15.8 | 14.7 | 11.9 | 16.8 | 19.9 | 16.9 | 15.5 | 18.3 | 19.2 | 18.3 | 22.0 |

**Table S5b.** Female mortality rates (per 100,000 population) by cause, age group and year in Scotland

| Age group | 2001 | 2002 | 2003 | 2004 | 2005 | 2006 | 2007 | 2008 | 2009 | 2010 | 2011 | 2012 | 2013 | 2014 | 2015 | 2016 | 2017 | 2018 | 2019 |
| --- | --- | --- | --- | --- | --- | --- | --- | --- | --- | --- | --- | --- | --- | --- | --- | --- | --- | --- | --- |
| All-Cause |  |  |  |  |  |  |  |  |  |  |  |  |  |  |  |  |  |  |  |
| 35-44 | 149.2 | 147.2 | 139.8 | 148.5 | 146.7 | 146.0 | 143.7 | 144.5 | 144.9 | 133.4 | 142.0 | 132.1 | 129.8 | 133.1 | 141.3 | 146.4 | 141.8 | 149.7 | 153.4 |
| 45-54 | 387.9 | 382.8 | 376.1 | 366.9 | 367.0 | 363.6 | 362.0 | 347.5 | 344.8 | 328.6 | 336.3 | 317.0 | 317.7 | 314.8 | 327.2 | 333.5 | 327.8 | 338.9 | 341.1 |
| 55-64 | 1014.3 | 1009.6 | 1019.0 | 934.6 | 932.2 | 919.3 | 909.7 | 865.6 | 838.1 | 835.8 | 810.4 | 794.0 | 778.8 | 748.0 | 766.4 | 766.3 | 754.3 | 769.4 | 759.9 |
| Combined 'Deaths of Despair' |  |  |  |  |  |  |  |  |  |  |  |  |  |  |  |  |  |  |  |
| 35-44 | 35.6 | 32.6 | 32.0 | 36.4 | 34.9 | 35.6 | 31.9 | 36.8 | 38.9 | 37.4 | 41.4 | 34.7 | 36.5 | 40.7 | 47.7 | 49.0 | 52.6 | 58.0 | 61.7 |
| 45-54 | 48.4 | 51.2 | 52.0 | 49.9 | 52.5 | 54.0 | 48.4 | 49.2 | 47.0 | 46.4 | 48.0 | 41.9 | 44.0 | 44.4 | 52.6 | 51.3 | 50.5 | 57.8 | 64.7 |
| 55-64 | 49.9 | 53.1 | 55.9 | 49.9 | 55.8 | 58.5 | 50.3 | 49.2 | 46.9 | 44.9 | 45.9 | 42.1 | 40.9 | 38.1 | 45.0 | 46.8 | 45.3 | 48.6 | 53.3 |
| Alcohol |  |  |  |  |  |  |  |  |  |  |  |  |  |  |  |  |  |  |  |
| 35-44 | 19.9 | 20.6 | 19.3 | 19.9 | 20.7 | 20.2 | 19.8 | 19.2 | 18.0 | 17.9 | 18.4 | 12.3 | 14.5 | 16.1 | 15.3 | 13.7 | 12.8 | 14.8 | 12.8 |
| 45-54 | 36.7 | 40.2 | 41.4 | 36.5 | 41.5 | 41.7 | 36.3 | 36.9 | 33.1 | 32.2 | 32.0 | 23.7 | 27.7 | 27.2 | 28.7 | 27.5 | 26.3 | 28.7 | 28.4 |
| 55-64 | 41.3 | 43.4 | 47.9 | 41.0 | 47.9 | 50.4 | 39.8 | 41.7 | 38.8 | 35.6 | 36.4 | 31.1 | 31.5 | 28.9 | 32.2 | 34.0 | 33.7 | 34.9 | 37.5 |
| Drugs |  |  |  |  |  |  |  |  |  |  |  |  |  |  |  |  |  |  |  |
| 35-44 | 4.8 | 3.7 | 4.7 | 5.6 | 5.7 | 7.8 | 5.0 | 8.5 | 13.0 | 10.5 | 14.6 | 13.8 | 15.4 | 15.3 | 23.2 | 26.5 | 33.1 | 34.0 | 40.2 |
| 45-54 | 2.8 | 2.7 | 3.5 | 3.5 | 3.4 | 4.4 | 4.8 | 5.5 | 6.6 | 7.5 | 9.0 | 10.4 | 9.8 | 9.6 | 14.3 | 15.2 | 17.2 | 20.9 | 26.1 |
| 55-64 | 2.0 | 2.2 | 2.2 | 2.7 | 1.8 | 2.4 | 4.1 | 3.4 | 2.4 | 4.7 | 4.6 | 5.7 | 4.1 | 3.9 | 5.4 | 5.8 | 5.5 | 7.5 | 9.0 |
| Suicide |  |  |  |  |  |  |  |  |  |  |  |  |  |  |  |  |  |  |  |
| 35-44 | 10.9 | 8.2 | 7.9 | 11.0 | 8.4 | 7.6 | 7.1 | 9.1 | 7.9 | 8.9 | 8.5 | 8.7 | 6.6 | 9.3 | 9.2 | 8.8 | 6.8 | 9.2 | 8.7 |
| 45-54 | 9.0 | 8.3 | 7.1 | 9.9 | 7.6 | 7.9 | 7.3 | 6.7 | 7.3 | 6.7 | 7.0 | 7.8 | 6.4 | 7.6 | 9.6 | 8.5 | 7.0 | 8.2 | 10.2 |
| 55-64 | 6.6 | 7.5 | 5.9 | 6.3 | 6.0 | 5.7 | 6.3 | 4.0 | 5.7 | 4.6 | 4.8 | 5.3 | 5.4 | 5.3 | 7.5 | 7.1 | 6.0 | 6.2 | 6.8 |

**Table S6a.**
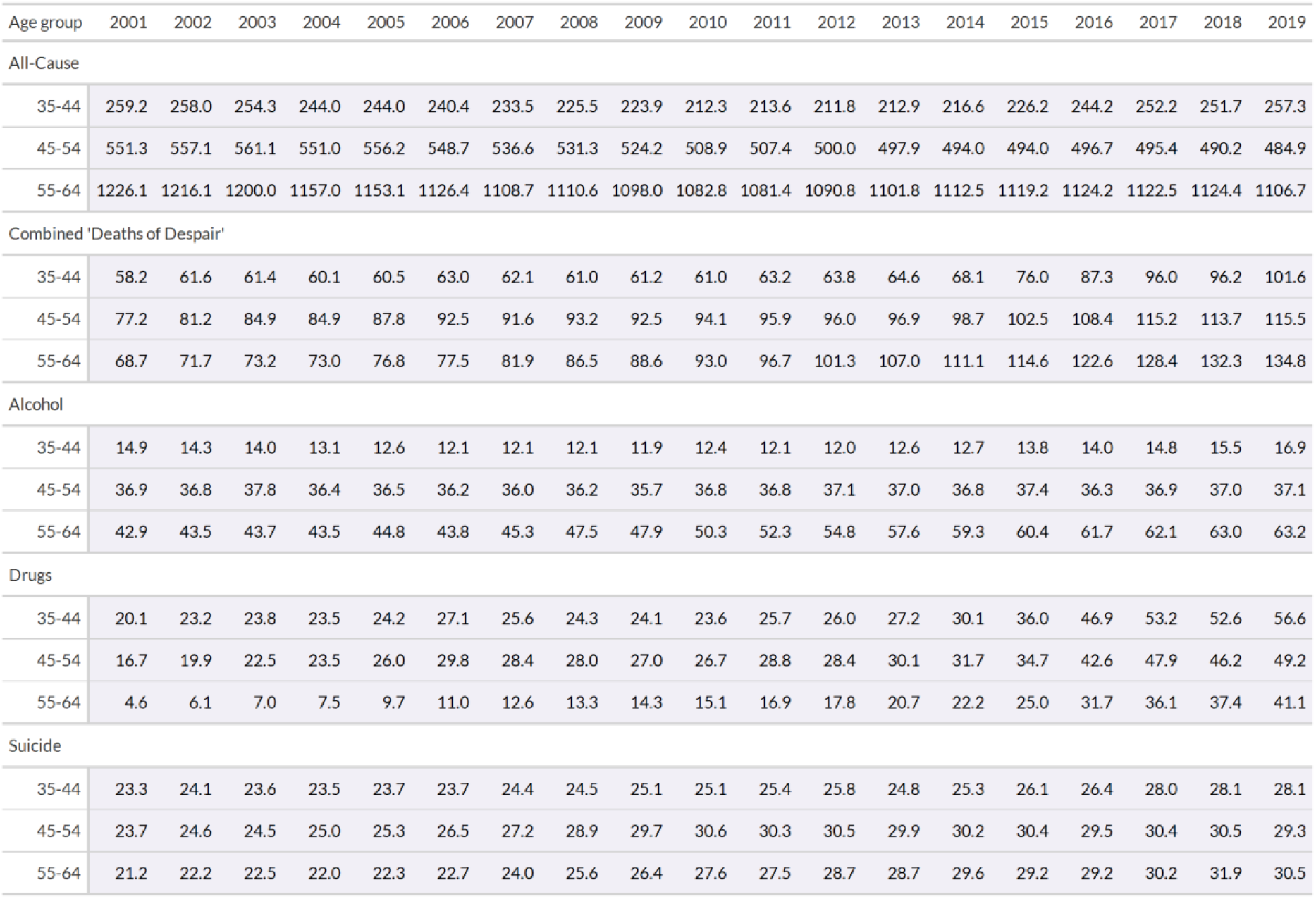
Male mortality rates (per 100,000 population) by cause, age group and year for the USA

| Age group | 2001 | 2002 | 2003 | 2004 | 2005 | 2006 | 2007 | 2008 | 2009 | 2010 | 2011 | 2012 | 2013 | 2014 | 2015 | 2016 | 2017 | 2018 | 2019 |
| --- | --- | --- | --- | --- | --- | --- | --- | --- | --- | --- | --- | --- | --- | --- | --- | --- | --- | --- | --- |
| All-Cause |  |  |  |  |  |  |  |  |  |  |  |  |  |  |  |  |  |  |  |
| 35-44 | 259.2 | 258.0 | 254.3 | 244.0 | 244.0 | 240.4 | 233.5 | 225.5 | 223.9 | 212.3 | 213.6 | 211.8 | 212.9 | 216.6 | 226.2 | 244.2 | 252.2 | 251.7 | 257.3 |
| 45-54 | 551.3 | 557.1 | 561.1 | 551.0 | 556.2 | 548.7 | 536.6 | 531.3 | 524.2 | 508.9 | 507.4 | 500.0 | 497.9 | 494.0 | 494.0 | 496.7 | 495.4 | 490.2 | 484.9 |
| 55-64 | 1226.1 | 1216.1 | 1200.0 | 1157.0 | 1153.1 | 1126.4 | 1108.7 | 1110.6 | 1098.0 | 1082.8 | 1081.4 | 1090.8 | 1101.8 | 1112.5 | 1119.2 | 1124.2 | 1122.5 | 1124.4 | 1106.7 |
| Combined 'Deaths of Despair' |  |  |  |  |  |  |  |  |  |  |  |  |  |  |  |  |  |  |  |
| 35-44 | 58.2 | 61.6 | 61.4 | 60.1 | 60.5 | 63.0 | 62.1 | 61.0 | 61.2 | 61.0 | 63.2 | 63.8 | 64.6 | 68.1 | 76.0 | 87.3 | 96.0 | 96.2 | 101.6 |
| 45-54 | 77.2 | 81.2 | 84.9 | 84.9 | 87.8 | 92.5 | 91.6 | 93.2 | 92.5 | 94.1 | 95.9 | 96.0 | 96.9 | 98.7 | 102.5 | 108.4 | 115.2 | 113.7 | 115.5 |
| 55-64 | 68.7 | 71.7 | 73.2 | 73.0 | 76.8 | 77.5 | 81.9 | 86.5 | 88.6 | 93.0 | 96.7 | 101.3 | 107.0 | 111.1 | 114.6 | 122.6 | 128.4 | 132.3 | 134.8 |
| Alcohol |  |  |  |  |  |  |  |  |  |  |  |  |  |  |  |  |  |  |  |
| 35-44 | 14.9 | 14.3 | 14.0 | 13.1 | 12.6 | 12.1 | 12.1 | 12.1 | 11.9 | 12.4 | 12.1 | 12.0 | 12.6 | 12.7 | 13.8 | 14.0 | 14.8 | 15.5 | 16.9 |
| 45-54 | 36.9 | 36.8 | 37.8 | 36.4 | 36.5 | 36.2 | 36.0 | 36.2 | 35.7 | 36.8 | 36.8 | 37.1 | 37.0 | 36.8 | 37.4 | 36.3 | 36.9 | 37.0 | 37.1 |
| 55-64 | 42.9 | 43.5 | 43.7 | 43.5 | 44.8 | 43.8 | 45.3 | 47.5 | 47.9 | 50.3 | 52.3 | 54.8 | 57.6 | 59.3 | 60.4 | 61.7 | 62.1 | 63.0 | 63.2 |
| Drugs |  |  |  |  |  |  |  |  |  |  |  |  |  |  |  |  |  |  |  |
| 35-44 | 20.1 | 23.2 | 23.8 | 23.5 | 24.2 | 27.1 | 25.6 | 24.3 | 24.1 | 23.6 | 25.7 | 26.0 | 27.2 | 30.1 | 36.0 | 46.9 | 53.2 | 52.6 | 56.6 |
| 45-54 | 16.7 | 19.9 | 22.5 | 23.5 | 26.0 | 29.8 | 28.4 | 28.0 | 27.0 | 26.7 | 28.8 | 28.4 | 30.1 | 31.7 | 34.7 | 42.6 | 47.9 | 46.2 | 49.2 |
| 55-64 | 4.6 | 6.1 | 7.0 | 7.5 | 9.7 | 11.0 | 12.6 | 13.3 | 14.3 | 15.1 | 16.9 | 17.8 | 20.7 | 22.2 | 25.0 | 31.7 | 36.1 | 37.4 | 41.1 |
| Suicide |  |  |  |  |  |  |  |  |  |  |  |  |  |  |  |  |  |  |  |
| 35-44 | 23.3 | 24.1 | 23.6 | 23.5 | 23.7 | 23.7 | 24.4 | 24.5 | 25.1 | 25.1 | 25.4 | 25.8 | 24.8 | 25.3 | 26.1 | 26.4 | 28.0 | 28.1 | 28.1 |
| 45-54 | 23.7 | 24.6 | 24.5 | 25.0 | 25.3 | 26.5 | 27.2 | 28.9 | 29.7 | 30.6 | 30.3 | 30.5 | 29.9 | 30.2 | 30.4 | 29.5 | 30.4 | 30.5 | 29.3 |
| 55-64 | 21.2 | 22.2 | 22.5 | 22.0 | 22.3 | 22.7 | 24.0 | 25.6 | 26.4 | 27.6 | 27.5 | 28.7 | 28.7 | 29.6 | 29.2 | 29.2 | 30.2 | 31.9 | 30.5 |

**Table S6b.**
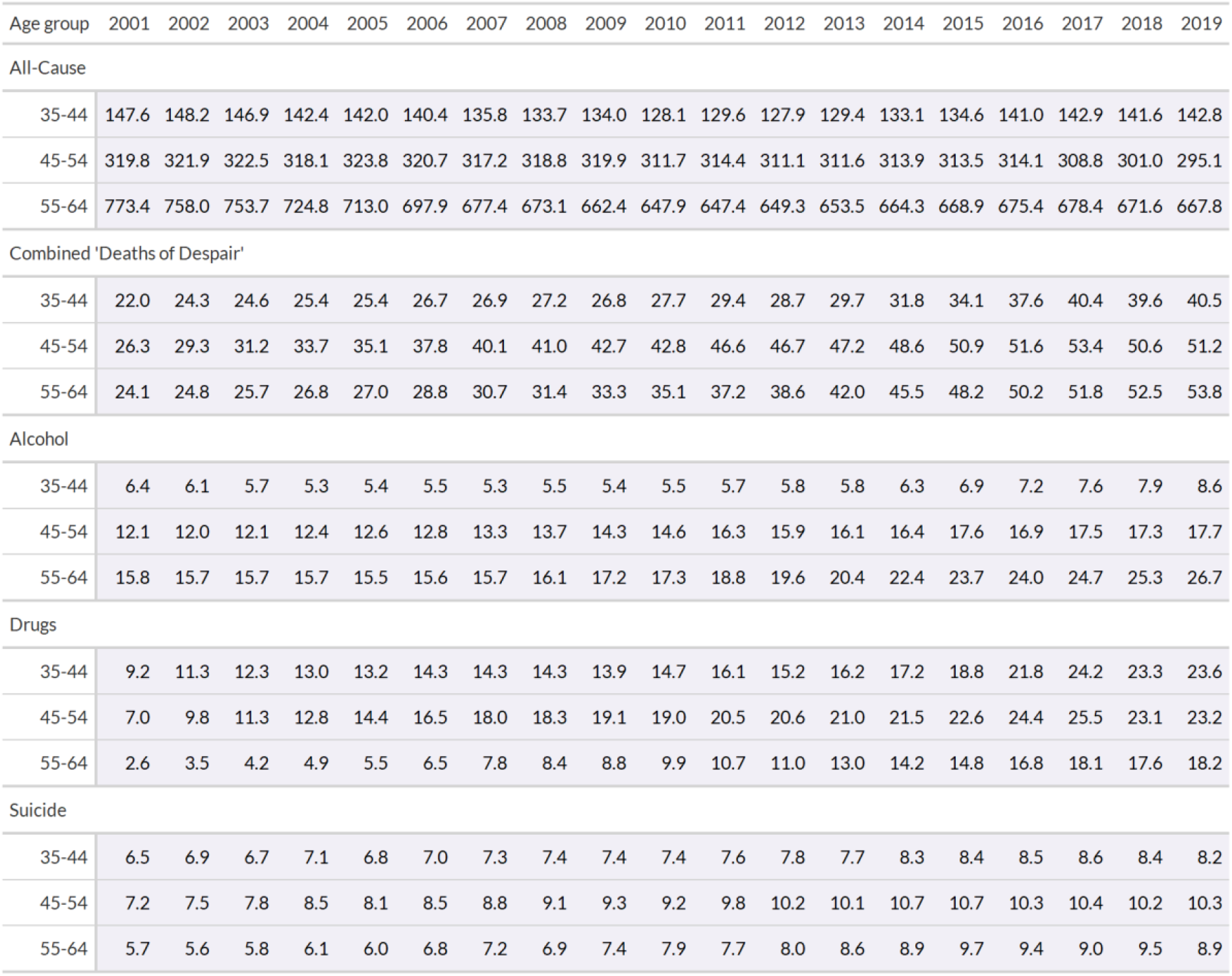
Female mortality rates (per 100,000 population) by cause, age group and year in the USA

## Footnotes

1 https://www.ons.gov.uk/peoplepopulationandcommunity/birthsdeathsandmarriages/deaths/bulletins/deathsrelatedtodrugpoisoninginenglandandwales/2020#drug-poisonings-from-selected-substances

2 https://www.ons.gov.uk/peoplepopulationandcommunity/birthsdeathsandmarriages/deaths/datasets/the21stcenturymortalityfilesdeathsdataset

3 https://www.nrscotland.gov.uk/statistics-and-data/statistics/statistics-by-theme/vital-events/general-publications/vital-events-reference-tables/archive

4 https://www.nisra.gov.uk/publications/registrar-general-annual-reports-2011-2019

5 Statistics Canada mortality data are accessible at: https://www150.statcan.gc.ca/t1/tbl1/en/cv.action?pid=1310039201

6 CDC Wonder data are accessible at: https://wonder.cdc.gov/.

7 https://www.nrscotland.gov.uk/files//statistics/drug-related-deaths/20/drug-related-deaths-20-pub.pdf

8 https://www.nrscotland.gov.uk/statistics-and-data/statistics/statistics-by-theme/vital-events/deaths/drug-related-deaths-in-scotland/2020

